# Interpretable Fine-tuned Large Language Models Facilitate Making Genetic Test Decisions for Rare Diseases

**DOI:** 10.64898/2026.02.26.26347223

**Authors:** Quan M. Nguyen, Fangyi Chen, Cong Liu, Ian M. Campbell, Gongbo Zhang, Da Wu, Katherine M. Szigety, Sarah E. Sheppard, Priyanka Ahimaz, Casey N. Ta, Wendy K. Chung, Chunhua Weng, Kai Wang

## Abstract

Clinical decision making often relies on expert judgment guided by established guidelines, which can be challenging to standardize and abstract to implement. For example, selecting between gene panels and whole exome/genome sequencing (WES/WGS) for rare disease diagnosis frequently requires interpretation of evidence-based recommendations from the American College of Medical Genetics and Genomics (ACMG) guideline. Traditional machine learning (ML) models predicting suitable genetic tests often face interpretability limitations. We hypothesize that large language models (LLMs) can be fine-tuned to “mimic” clinicians’ reasoning patterns by interpreting and applying clinical guidelines with chain-of-thought (CoT). We present RareDAI, an integrative approach that addresses this challenge by analyzing heterogeneous clinical data, including unstructured notes and structured Phecodes. Using seven domain-specific questions, we guide the Llama 3.1 and Qwen 3 models to generate structured CoT outputs. These outputs are refined via our proposed self-distillation fine-tuning (SDFT) approach, enabling the model to produce interpretable reasoning prior to recommendation. RareDAI outperforms traditional supervised fine-tuning and base LLMs (e.g., Llama 3.1, GPT-4) by up to 10-20% in all metrics (accuracy, precision, recall, and F1-score) on both in-house data and external data, effectively assisting clinicians in selecting between diagnostic modalities across healthcare systems.

## Introduction

Clinical genetic testing guidelines, including those released by the American College of Medical Genetics and Genomics (ACMG), are intended make the provision of genetic testing more accurate, timely, cost-effective, and equitable^1^. Nonetheless, implementation of guidelines into real-world clinical practice often lags substantially from publication, even when evidence is strong^2,3^. This phenomenon is exacerbated when complex and nuanced guidelines collide with the reality of busy clinical practice. One potential avenue to improve guideline adherence is clinical decision support (CDS) systems^4^. However, to provide effective support, such a system must accurately translate guidelines into machine interpretable code.

Some guidelines, such as the use of fasting glucose levels to determine risk of diabetes, are relatively straightforward to implement. However, others, such as the ACMG testing guidelines for genetic testing for patients with congenital anomalies^1^ can be more complex and abstract. Whole exome or genome sequencing are recommended by the ACMG as first- or second-tier diagnostic tools for pediatric patients with congenital anomalies, developmental delays, or intellectual disabilities^1^. This creates a critical challenge in translating this evidence based, yet somewhat vague guidelines into concrete actions for individual patients. Traditional machine learning models rely on predefined structured features, limiting their ability to capture extensive temporal context and complex guideline logic. However, large language models (LLMs) are designed to model contextual relationships in free text and have shown strong performance on long-form clinical text and guideline-based reasoning tasks^5–8^, though their reliability in medical decision-making remains an open question.

The challenge of improving the provision of molecular genetic testing is essential as such testing is critical to the diagnosis of rare diseases affecting over 25 million people in the United States^9^ and approximately 300 million people worldwide^10^. This process can often be lengthy and complex^11–13^. Selecting the right testing strategy early is crucial to limit delay in diagnosis and cost. Currently, several primary types of molecular genetic tests—single-gene tests, gene panels, and exome/genome sequencing—are commonly used, each with distinct advantages and limitations^14,15^. Single-gene tests are often preferred for well-defined Mendelian disorders such as myotonic dystrophy due to speed, specialized testing requirements, or affordability^14–16^. However, when signs and symptoms indicate the differential diagnosis is broad or when a single-gene test is inconclusive, a gene panel may be preferable^14^. For example, depending on the clinical setting, gene panels for epilepsy may screen between 70 and 377 different genes, enhancing the likelihood of identifying the causative genes at a reasonable cost^15^. Exome or genome sequencing provide the most comprehensive analysis but are typically reserved for cases with nonspecific symptoms, a large or diverse of genes in the differential, particular urgency, or prior inconclusive testing^14,15^. Despite declining sequencing costs, choosing the optimal genetic test remains difficult and suboptimal decisions can delay diagnosis, increase stress and costs, and lead to inappropriate treatment or irreversible disease progression when effective therapies are available.

The recent development of LLMs has led to a wide range of biomedical applications^7,17–22^, such as generating clinical reports, answering medical questions, assisting in research, improving patient communication, and streamlining administrative tasks. LLMs can often answer questions, summarize, paraphrase and translate text on a level that is sometimes indistinguishable from human capabilities^23^. The Transformer architecture^24^ has been the basis for some of the most advanced LLMs currently in use, such as GPT-4^25^, Llama 3.1^26^, BERT^5^, and T5^27^. For example, a variety of Transformer-based models, including PhenoGPT^7^, PubMedBERT^22^, BioBERT^6^, and Bioformer^17^, have demonstrated remarkable potential in processing complex biomedical texts and extracting phenotypic insights, while Google’s MedPalm^28^ specializes in delivering accurate responses to medical questions. Therefore, integrating LLMs into clinical workflow may play a future role in clinical decision making, including the field of clinical genetics.

The most suitable genetic testing strategy for patients with rare diseases is determined by genetic counselors or physicians through clinical evaluation^15^. However, recent work suggests that machine learning (ML) techniques^29,30^ show promise in selecting appropriate testing. With current technology, clinical evaluation likely remains most accurate. However, diagnoses may be delayed due to barriers to accessing care, exacerbated by a shortage of genetic specialists^31^. Furthermore, clinical judgment from human experts is shaped by years of experience, making it difficult for less experienced clinicians to replicate. Hence, harnessing the power of computational algorithms to develop automated decision-making methods may address these challenges. Conventional machine learning approaches typically rely on structured inputs such as billing codes, laboratory results, or curated phenotype features. While these data sources capture important clinical signals, a substantial proportion of clinically relevant information remains embedded in unstructured clinical notes, which is not directly accessible to such models without additional feature engineering or text processing. As a result, relationships expressed in free-text narratives may be incompletely represented. Furthermore, unstructured clinical notes are often lengthy and disorganized, posing challenges for their reliable conversion into structured feature sets despite recent efforts^7,29,30,32^. This is an area where LLMs perform well, given their ability to comprehend context, intelligently capture the semantic nuances of textual patterns, providing reasoned answers and guiding the decision-making process.

This study proposes a novel LLM-based approach, referred to as RareDAI, to assist clinicians in determining diagnostic test choice - gene panels versus exome/genome sequencing (referred to as genome sequencing hereafter) - by analyzing recorded human phenotype ontologies (HPO^33^), disease diagnosis codes (ICD-10), and clinical notes. Given that gene panels and next-generation sequencing (NGS) tests have become the preferred options over single-gene tests, we are specifically interested in determining which patients are best suited for these two methods of genetic testing. Previous research has shown that phenotypes and EHR billing codes offer valuable insights into identifying genetic diseases^34–36^, which could also aid in selecting the appropriate genetic testing method^37^. Additionally, the chain-of-thought (CoT) approach has significantly enhanced LLM performance, as demonstrated by numerous recent studies^38–41^. In the implementation of RareDAI, we leverage base LLMs to generate detailed CoT explanations, which are then integrated into the fine-tuning process. Additionally, to further examine the performance of RareDAI, we also performed a thorough comparison of the impact of incorporating phenotypes, diagnosis codes, and self-distilled CoT, alongside varying LLM sizes.

## Results

### Overview of the Fine-Tuning Strategy and Experimental Methods

We developed an LLM-based method to assist providers in selecting an appropriate genetic testing strategy based on information documented in clinical notes. This method, we call RareDAI, utilizes a self-distillation fine-tuning strategy to imitate the reasoning process of an expert clinician. An illustration of the fine-tuning strategy employed in RareDAI is shown in **Figure 1**. Compared to conventional supervised fine-tuning, RareDAI requires two separate steps. In step one, a larger, higher performance base model (e.g., Llama-3.1-70B or Qwen3-30B-A3B-Thinking-2507) is guided to answers seven expert- and guideline-derived questions based on clinical notes and test labels (panel vs. exome/genome), creating a “distilled” dataset. In step two, a smaller model (Llama-3.1-8B or Qwen3-8B) is fine-tuned on this dataset to learn how to make informed test recommendations with detailed explanations. We additionally summarized clinical notes prior to Step 1 and trained and evaluated these models alongside models using raw notes.

**Figure 1.**
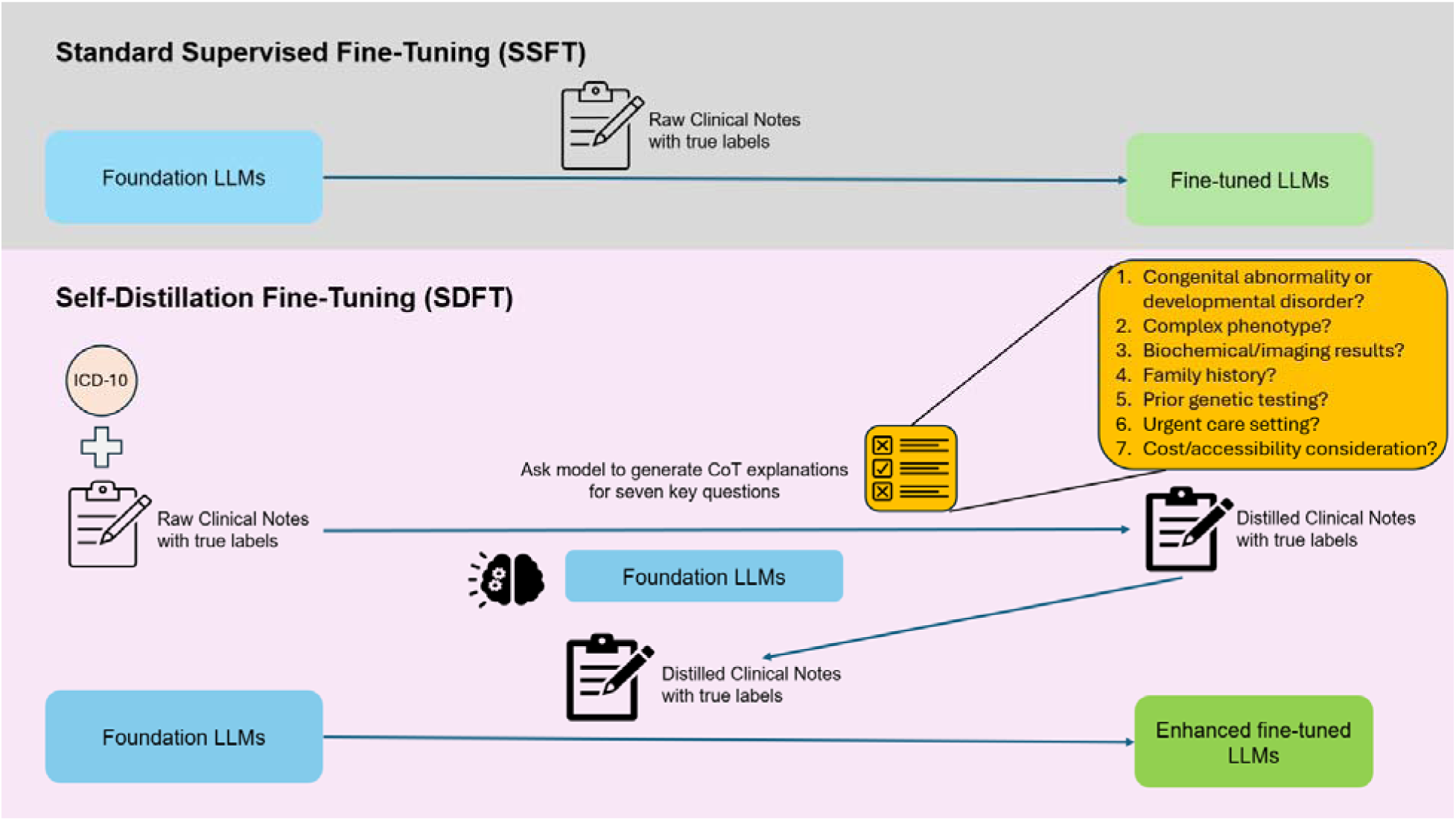
Illustration of the self-distillation fine-tuning strategy implemented in RareDAI. Seven questions are fully presented in Supplementary Figure 2.

### Evaluation on Base Models (without fine-tuning)

We first evaluate the performance of “off-the-shelf” base LLMs with different parameter sizes (8B and 70B) under various conditions, to establish the baseline performance. These models are referred to as base models, since they are not subject to fine-tuning process. However performance can vary depending on model size, quantization strategy and prompting tokens. Quantization reduces the precision of model parameters to decrease total memory requirements and computation time while sacrificing performance. We evaluated the models on 127 patients who underwent genetic testing, including 77 with exome/genome sequencing and 50 with gene panels. Using both unstructured clinical notes and structured information, the 70B base model with 4-bit quantization achieved 51% accuracy and a 55% F1-score. Meanwhile, the 70B base model with 16-bit precision (the default) reached 65% accuracy and a 62% F1-score. The 8B base model (16-bit) performed similarly with 64% accuracy and a 61% F1-score (Table 1). In comparison, when given seven structured questions in the prompting procedure, the 70B model showed some improvement, with a 3% increase in F1-score for both 4bit and 16bit versions (**Supplementary Table 1**), indicating the guidelines help improve reasoning. However, the 8B model’s performance declined slightly under the same conditions, suggesting that it struggles with additional contextual input.

**Table 1:**
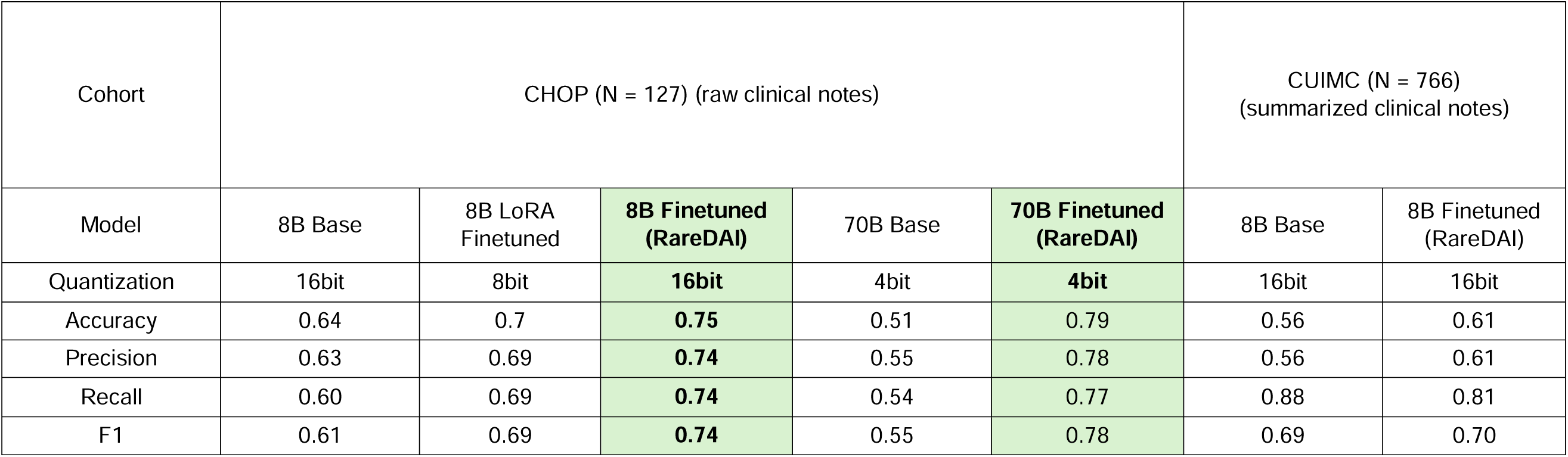
Performance of Fine-Tuned (RareDAI) vs. Base Models on CHOP and CUIMC Test Datasets. Our best performing fine-tuned models (8B with CoT/ICD and 70B with HPO/CoT/ICD) outperform foundation Llama-3.1-8B and 70B models (base), with supervised fine-tuning on domain-specific data improving task performance and expert alignment. Quantized models (8B LoRA) underperform 16-bit models, likely due to information loss. Fine-tuned RareDAI achieves slightly higher accuracy, precision, and F1-score on the Columbia dataset.

### Evaluation on Fine-tuned Models that Incorporates Critical Thinking

The goal of the fine-tuning strategy (**Figure 1**) is to elicit structured reasoning across seven specific questions, before deriving the final recommendations. We hypothesized that the answers to these questions would server as human-understandable and interpretable explanations of the final recommendation. **Table 1** shows a 12-26% increase in all metrics for both the fine-tuned 8B and 70B models compared to their corresponding base models. The 16-bit precision fine-tuned 8B model achieved 74% on average across all metrics compared to under 65% in its base form, while the fine-tuned 70B model achieved roughly 79% across all metrics.

We compared RareDAI to a private version of ChatGPT 5.0 Enterprise^53^ utilized at the Children’s Hospital of Philadelphia (CHOP), on the same test dataset (N = 127). ChatGPT@CHOP allows users to securely use confidential data and ensure patient privacy, while providing the same power as the standard ChatGPT 5.0 model. This experiment was performed manually, since API access to ChatGPT@CHOP was not available. While ChatGPT has demonstrated value in explaining complex topics and generating ideas or performing reasonably on medical exam questions due to its sophisticated architecture and extensive training^54–57^, it is likely that specialized medical topics (such as genetic testing) require advanced domain expertise for more accurate interpretation. Therefore, without tailored training on medical corpora, ChatGPT may underperform in such areas. This hypothesis is supported by **Figure 2**, which shows ChatGPT achieving only 57% accuracy, 61% recall, and 62% for both precision and F1-score in predicting genetic tests, falling short of RareDAI’s performance.

**Figure 2.**
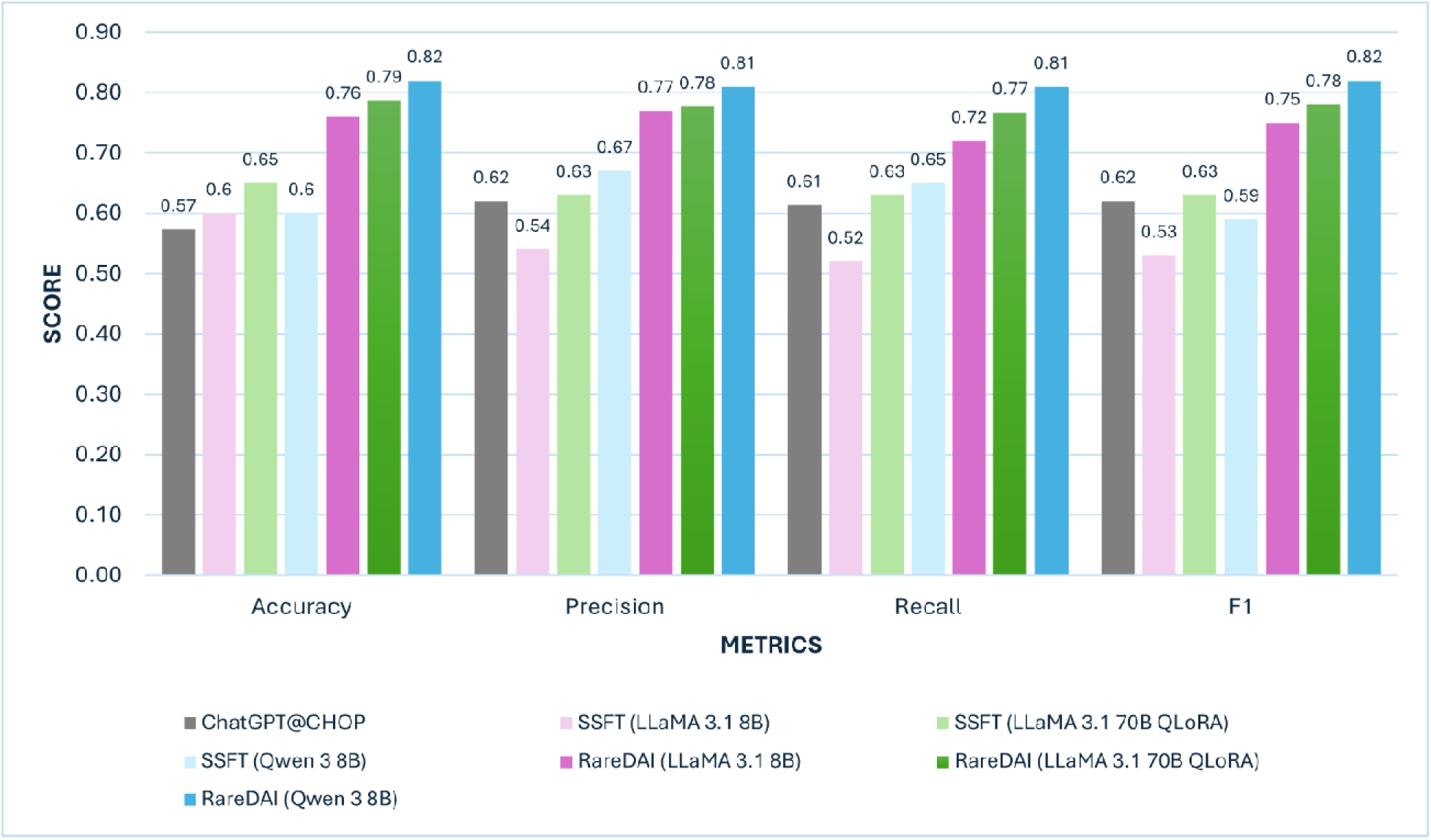
Performance of RareDAI (Llama-3.1-8B, 70B models and Qwen3-8B) on the raw full CHOP clinical notes. Our top-performing LLMs outperform ChatGPT@CHOP (a private ChatGPT Enterprise model for the Children’s Hospital of Philadelphia) and Standard Supervised Fine-tuning (SSFT) methods. Its superior results are attributed to its advanced ability to capture nuanced textual representations, fine-tuned on a specialized task, and employ chain-of-thought reasoning, enabling more accurate predictions.

### Additional Evaluation on Qwen3 8B

Qwen3 has demonstrated strong performance across a wide range of tasks, including medical applications^50,58,59^. In this study, we further evaluated Qwen3 alongside Llama models to assess the robustness of our approach across different model architectures. We followed the same pipeline used for Llama but employed Qwen3-30B-A3B-Thinking-2507 for CoT generation and fine-tuned Qwen3 8B as the inference model. Overall, the fine-tuned Qwen3 8B model used with the base model’s pre-trained reasoning enabled (specified enable_thinking = True) achieved slightly higher performance than the fine-tuned Llama-3.1-8B model across most modeling strategies on raw clinical notes (**Table 2**). When operating in thinking mode, the model first generates intermediate reasoning within <think> and </think> tokens, where it aggregates relevant information, before producing the final response^50^. Under the standard supervised fine-tuning setting without CoT, Qwen3 8B reached 60% accuracy, compared with 60% for Llama-3.1-8B, though both underperformed Llama 70B (65%) (**Figure 2**). When CoT and the Qwen-specific thinking mechanism were incorporated, Qwen3 8B demonstrated substantial gains and, in several configurations, outperformed both Llama models. The best-performing Qwen3 configuration (HPO–ICD–CoT–Thinking) on long clinical notes achieved 82% accuracy and an F1 score of 81%.

**Table 2:** Performance on Long Clinical Notes Across Models and Prompting Strategies. This table reports Accuracy, Precision, Recall, and F1 scores (with 95% bootstrap confidence intervals) for baseline (standard supervised fine-tuning) and Rare-DAI Llama and Qwen models evaluated on long clinical notes under different prompting configurations, including baseline, chain-of-thought, and structured phenotype-aware prompts. Top performing models are Llama-70B (with HPO+ICD+CoT) and Qwen3-8B-Thinking (with HPO+ICD+CoT).

| Prompting Strategy | Model | Accuracy | Precision | Recall | F1 |
| --- | --- | --- | --- | --- | --- |
| **Baseline (SSFT)** | Llama-8B | 0.60 (0.51-0.69) | 0.54 (0.41-0.69) | 0.52 (0.46-0.58) | 0.53 (0.43-0.63) |
|  | Llama-70B | 0.65 (0.57-0.73) | 0.63 (0.54-0.72) | 0.63 (0.54-0.72) | 0.63 (0.57-0.74) |
|  | Qwen3-8B | 0.60 (0.51-0.69) | 0.67 (0.59-0.75) | 0.65 (0.57-0.72) | 0.59 (0.50-0.68) |
| **CoT Only** | Llama-8B | 0.76 (0.69-0.84) | 0.77 (0.69-0.86) | 0.72 (0.65-0.81) | 0.75 (0.67-0.84) |
|  | Llama-70B | 0.72 (0.65-0.80) | 0.71 (0.63-0.79) | 0.71 (0.63-0.79) | 0.72 (0.65-0.79) |
|  | Qwen3-8B | 0.69 (0.61-0.76) | 0.69 (0.61-0.76) | 0.69 (0.62-0.78) | 0.69 (0.61-0.77) |
|  | Qwen3-8B-Thinking | 0.74 (0.66-0.82) | 0.73 (0.64-0.81) | 0.72 (0.64-0.81) | 0.74 (0.66-0.82) |
| **ICD+CoT** | Llama-8B | 0.75 (0.69-0.84) | 0.74 (0.67-0.83) | 0.74 (0.67-0.82) | 0.74 (0.68-0.83) |
|  | Llama-70B | 0.76 (0.69-0.84) | 0.76 (0.68-0.83) | 0.74 (0.66-0.82) | 0.76 (0.68-0.83) |
|  | Qwen3-8B | 0.74 (0.66-0.81) | 0.74 (0.66-0.81) | 0.75 (0.70-0.82) | 0.74 (0.67-0.81) |
|  | Qwen3-8B-Thinking | 0.78 (0.68-0.83) | 0.77 (0.67-0.82) | 0.77 (0.67-0.82) | 0.78 (0.68-0.83) |
| **HPO+ICD+CoT** | Llama-8B | 0.75 (0.68-0.83) | 0.74 (0.66-0.82) | 0.72 (0.64-0.81) | 0.74 (0.67-0.82) |
|  | **Llama-70B** | **0.79 (0.72-0.87)** | **0.78 (0.71-0.86)** | **0.77 (0.70-0.86)** | **0.78 (0.72-0.87)** |
|  | Qwen3-8B | 0.71 (0.63-0.79) | 0.70 (0.62-0.78) | 0.71 (0.63-0.79) | 0.71 (0.63-0.79) |
|  | **Qwen3-8B-Thinking** | **0.82 (0.75-0.89)** | **0.81 (0.74-0.88)** | **0.81 (0.73-0.88)** | **0.81 (0.75-0.90)** |

### Summarized Structured Notes Improve CoT-Based Reasoning

Because manual verification of all summarized notes across the training, validation, and test sets would be impractical, we conducted a targeted manual review of 30 randomly selected summaries from the test set. We confirmed that the generated summaries accurately reflected necessary information present in the original clinical notes and appropriately addressed the prompted questions, with no hallucinated clinical facts observed.

We evaluated four modeling strategies—baseline supervised fine-tuning without CoT, CoT-only, ICD–CoT, and HPO–ICD–CoT—across three model backbones: Llama 8B (16-bit), Llama 70B (4-bit), and Qwen3 8B (16-bit). Overall, models trained on summarized structured notes consistently outperformed their counterparts trained on raw clinical notes, with accuracy improvements of several percentage points across most settings. In particular, Llama 8B and Qwen3 8B trained with ICD/CoT achieved accuracies of 86% and 84%, respectively, when trained on summarized notes, compared to 75% and 74% when trained on raw notes.

Finally, incorporating structured phenotype information improved performance only when paired with models capable of strong reasoning. Qwen3-8B with an explicit thinking process and Llama-70B both showed accuracy gains when HPO features were included (82% and 79% in accuracy, respectively) compared with runs without HPO information (78% and 76%). In contrast, models with weaker reasoning exhibited limited benefit, indicating that structured phenotype representations are most effective when the model can explicitly reason over them in the presence of noisy clinical data.

### Highlighted Examples and Analysis of Successful Assessments by RareDAI

We next examined two successful examples of recommending genome sequencing (**Figure 4**) and gene panel (**Figure 5**) by our top performing model (fine-tuned Qwen3-8B CoT/ICD). While large-scale manual evaluation of every reasoning step was not feasible due to limited expert availability, these two representative cases illustrate the broader effectiveness of RareDAI: its reasoning consistently aligns with clinical guidelines and captures relevant patient details. These two examples are synthetic and were manually compiled based on summarized representations of the original clinical notes. All personal health information was removed or replaced, and only sufficient clinical details were retained to support chain-of-thought evaluation. In the first case (**Figure 4**), the infant exhibits severe growth failure and feeding difficulty in the absence of distinctive dysmorphic features or organ-specific findings, constituting a nonspecific developmental presentation. According to ACMG guidelines, genome-wide sequencing (exome/genome) is indicated as a first-tier diagnostic modality for unexplained growth failure, particularly given the noncontributory family history, normal newborn screen, and lack of prior genetic testing. Given the broad differential spanning numerous genetic etiologies, targeted panels are susceptible to false negatives and diagnostic delay; thus, comprehensive genome sequencing represents the most appropriate initial test to optimize diagnostic yield and expedite management.

In the second case (**Figure 5**), a targeted hearing-loss gene panel is recommended as the first-line test. The child has isolated, childhood-onset bilateral sensorineural hearing loss with normal growth, development, and neurologic and otologic examinations. MRI shows no structural abnormalities, and there are no syndromic features or multisystem involvement. Family history is limited to a first cousin with hearing loss attributed to congenital CMV, which does not suggest a clear inherited pattern. Consistent with ACMG guidance, phenotype-directed panel testing is preferred in this setting, offering faster turnaround, lower cost, and fewer incidental findings than exome or genome sequencing. Broader genomic testing would be reserved for cases with negative panel results, emerging syndromic features, or clearer evidence of familial inheritance. Our clinical expert confirmed that this reasoning was appropriate.

Both examples reflect the definitions given in our instruction prompt and use available information from the input to provide detailed explanations. We observe that in both cases, the models attempted to connect critical details from the input and offered reasoning and explanations to justify why one genetic test may be considered better than the other (**Figure 3**). By offering step-by-step analysis, models have a better chance of generating correct answers, as each generated token is based on the prior context. Therefore, with persuasive chain-of-thought reasoning, LLMs are better able to predict the final assessment.

**Figure 3.**
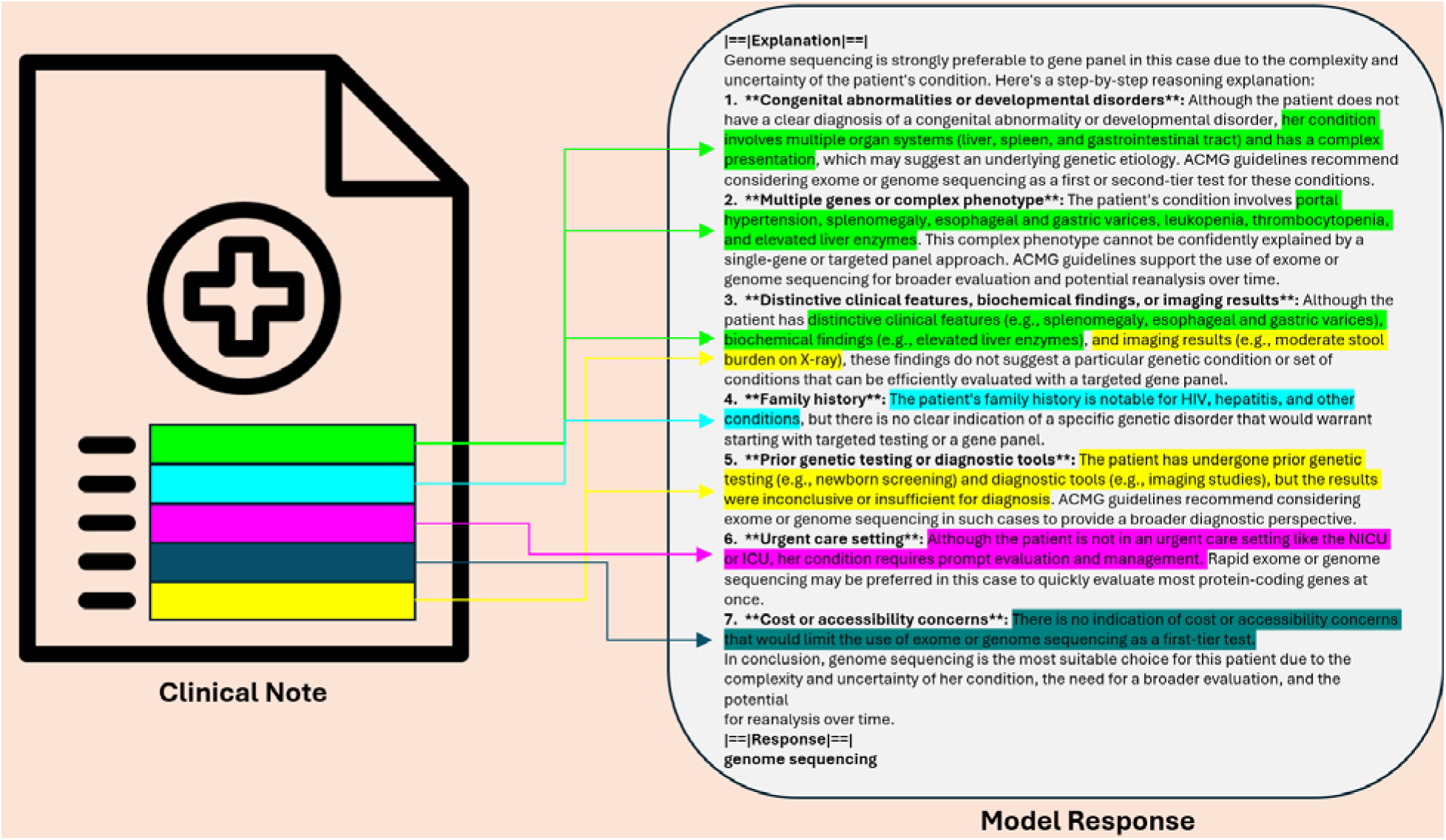
Case Study. Identical information is color-coded, allowing direct comparison between the input clinical note and model response. The same color represents the same information, demonstrating that the model effectively utilizes key clinical details for inference. This structured approach ensures that genetic testing recommendations are evidence-based and clinically relevant. See Supplementary Figures S5 and S6 for details.

We also showcased the responses from our best Llama-3.1-8B (ICD/CoT trained on summarized notes) (**Supplementary Figure 5**), ChatGPT (**Supplementary Figure 6**), and the Llama-3.1-8B base model (**Supplementary Figure 7**) for the first patient case (true label: genome sequencing; see **Figure 4**). Our RareDAI-Llama 8B’s response is also well-reasoned and reaches an appropriate conclusion (genome sequencing) for a nonspecific presentation. It highlights key considerations—broad differential, noncontributory family history, lack of prior genetic testing, and the inefficiency of panels in the absence of a targeted syndrome—which align with ACMG guidance favoring genome-wide testing. However, some justifications are less precise (e.g., citing “distinctive features” like thin appearance, implying urgency outside critical care, and overemphasizing “rapid” turnaround without context) and could be strengthened by explicitly framing the phenotype as nonspecific developmental/growth failure and directly tying the recommendation to first-tier ACMG indications. Overall, it is a solid rationale, though it could be tightened to avoid minor inconsistencies and better anchor claims to guideline language. Meanwhile, ChatGPT incorrectly recommended a gene panel because it ignores ACMG first-tier recommendations for nonspecific, severe growth failure by favoring a narrow panel and restricts testing to selected malabsorption/metabolic genes, increasing risk of missed diagnoses across the broad differential, whereas the RareDAI-Qwen3 correctly recommends genome sequencing by using concrete clinical metrics and guideline mapping. On the other hand, the Llama-3.1-8B base model misapplies ACMG guidance by favoring a panel despite a nonspecific, severe growth failure presentation where genome/exome is recommended first-tier. It repeats unsupported assumptions (e.g., “distinctive features,” family-history-driven testing”, “newborn screen sufficiency”) and introduces contradictions.

**Figure 4:**
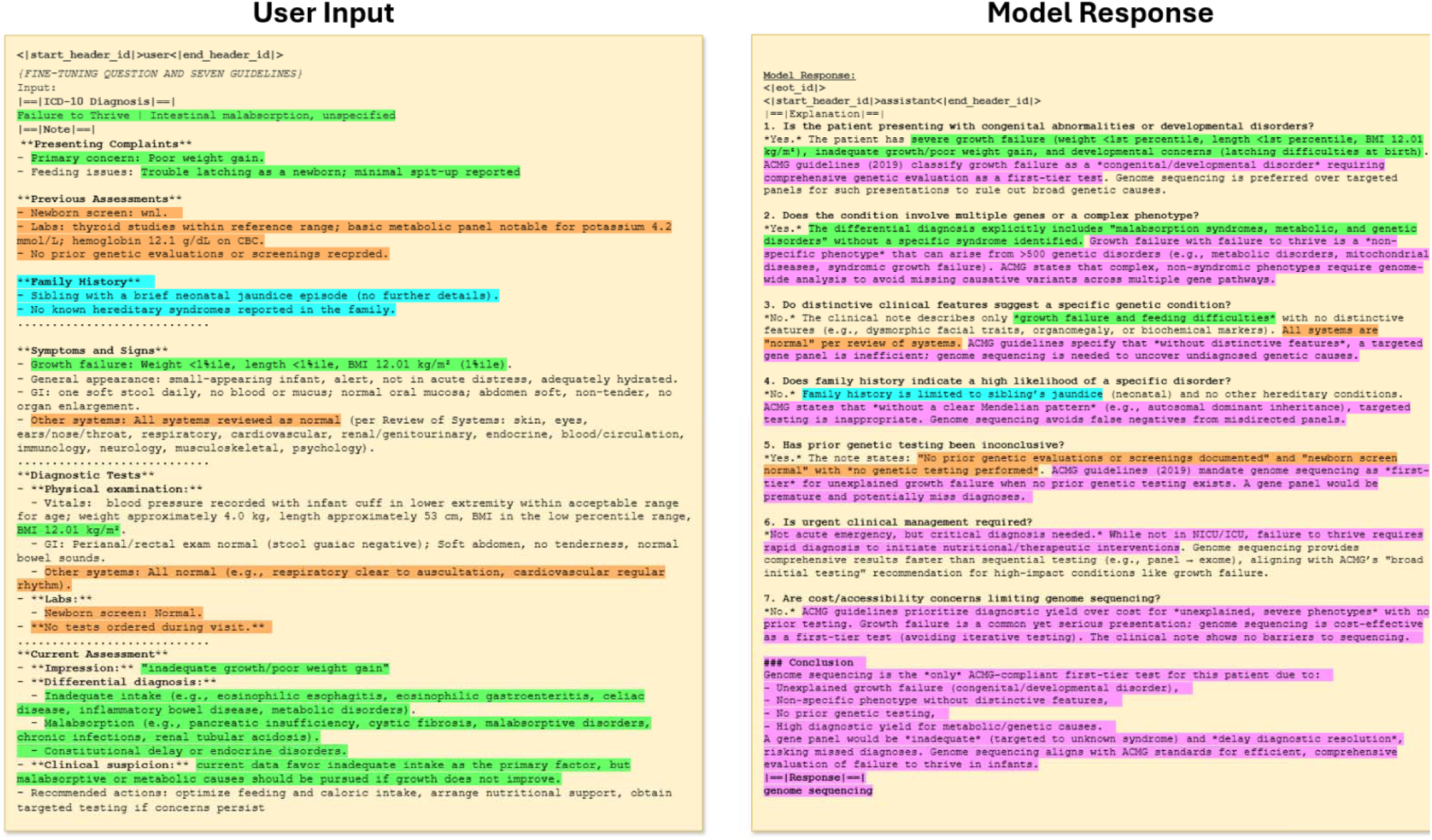
An example of a Genome Sequencing test case. Identical information is color-coded, allowing direct comparison between the input and model response. The same color represents the same information, demonstrating that the model effectively utilizes key clinical details for inference. Green: phenotypic information, Orange: prior testing results, Aqua: family history, Violet: Model reasoning. This structured approach ensures that genetic testing recommendations are evidence-based and clinically relevant. Model used: fine-tuned Qwen3-8B (16bit) CoT/ ICD on summarized notes. The clinical note shown below is a synthetic example designed to simulate a summarized real-world test case.

**Figure 5:**
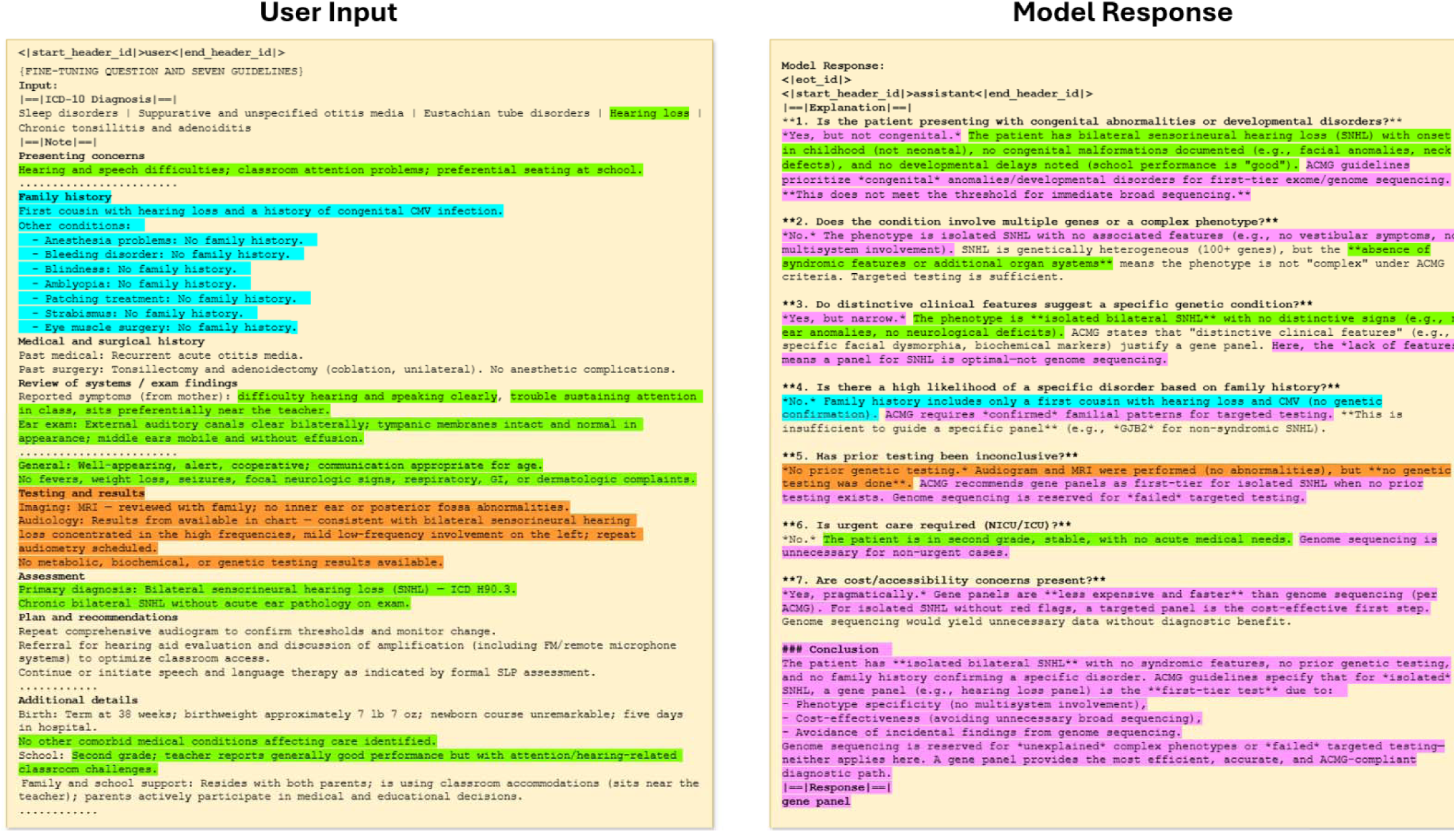
An example of a Gene Panel test case. Identical information is color-coded, allowing direct comparison between the input and model response. The same color represents the same information, demonstrating that the model effectively utilizes key clinical details for inference. Green: phenotypic information, Orange: prior testing results, Aqua: family history, Violet: Model reasoning. This structured approach ensures that genetic testing recommendations are evidence-based and clinically relevant. Model used: fine-tuned Qwen3-8B (16bit) CoT/ ICD on summarized notes. The clinical note shown below is a synthetic example designed to simulate a summarized real-world test case.

### External Validation on an Independent Dataset on the Robustness of the Models

We further evaluated the performance of base Llama-3.1-8B and our RareDAI-Llama 8B model trained on summarized clinical notes on an independent external dataset from Columbia University Irving Medical Center (N = 766), primarily on children under 10 years old from diverse backgrounds. To reduce variability across institutions and to prevent information leakage, the model trained on summarized versions of clinical notes was used for external validation. The 8B base model achieved only 56% accuracy and 69% F1 score. In contrast, RareDAI achieved 61% accuracy and 70% F1 score. While the performance metrics are lower compared to our in-house data, the results highlight the advantages of fine-tuned LLMs over base models. The lower performance may be due to differences in documentation styles and structures between medical institutions or differences in how clinicians interpret the original guidelines to make clinical decisions.

### Model diagnostics: the impact of Chain-of-Thought and seven key questions

We next performed model diagnostics to understand what factors drive the improved performance over base models. A key innovation of RareDAI was the use of CoT reasoning generated by base LLMs to perform self-distillation fine-tuning, which subsequently improves performance in this classification task. We found that incorporating CoT indeed significantly enhanced performance (**Tables 2 and 3**). Incorporating structured questions significantly improves model performance by directing attention to critical pieces of information within lengthy clinical notes. Without these targeted questions and CoT, the 70B model (standard supervised fine-tuning) achieved ∼63% in all metrics on average. However, when the model was guided by seven key questions clinical questions, its performance surged to nearly 79% across all evaluation metrics. Additionally, as previously described, even the base model (70B, 16bit) performs better (76% for precision and 65% for F1-score) when providing these questions in the prompt compared to simply asking for a recommendation (67% and 62%, respectively) (**Supplementary Table 1**). These improvements demonstrate the power of structured reasoning in LLMs—by explicitly aligning the model’s focus with established clinical guidelines for determining appropriate genetic tests, we enhance both accuracy and reliability. These questions serve as anchors, ensuring that the model systematically evaluates the most relevant clinical factors rather than relying solely on the next token prediction based on its pre-trained corpus. This approach highlights the importance of domain-specific prompting in medical AI, reinforcing that even large models benefit from structured decision-making frameworks based on expert knowledge.

**Table 3:** Performance on Summarized Clinical Notes Across Models and Prompting Strategies. This table reports Accuracy, Precision, Recall, and F1 scores (with 95% bootstrap confidence intervals) for baseline (standard supervised fine-tuning) and Rare-DAI Llama and Qwen models evaluated on long clinical notes under different prompting configurations, including baseline, chain-of-thought, and structured phenotype-aware prompts. Top performing models are Llama-8B (with ICD+CoT) and Qwen3-8B (with ICD+CoT).

| Prompting Strategy | Model | Accuracy | Precision | Recall | F1 |
| --- | --- | --- | --- | --- | --- |
| Baseline (SSFT) | Llama-8B | 0.66 (0.58-0.75) | 0.70 (0.63-0.78) | 0.70 (0.63-0.77) | 0.66 (0.58-0.74) |
|  | Llama-70B | - | - | - | - |
|  | Qwen3-8B | 0.70 (0.63-0.79) | 0.69 (0.61-0.78) | 0.68 (0.61-0.77) | 0.70 (0.62-0.79) |
| CoT Only | Llama-8B | 0.79 (0.73-0.87) | 0.78 (0.72-0.87) | 0.77 (0.72-0.86) | 0.79 (0.73-0.87) |
|  | Llama-70B | - | - | - | - |
|  | Qwen3-8B | 0.74 (0.66-0.82) | 0.73 (0.66-0.81) | 0.74 (0.66-0.82) | 0.74 (0.66-0.82) |
|  | Qwen3-8B-Thinking | 0.72 (0.65-0.80) | 0.74 (0.66-0.80) | 0.74 (0.67-0.81) | 0.73 (0.65-0.80) |
| ICD+CoT | **Llama-8B** | **0.86 (0.80-0.91)** | **0.87 (0.80-0.93)** | **0.83 (0.76-0.90)** | **0.85 (0.79-0.91)** |
|  | Llama-70B | 0.79 (0.71-0.86) | 0.78 (0.70-0.85) | 0.79 (0.72-0.86) | 0.79 (0.71-0.86) |
|  | **Qwen3-8B** | **0.84 (0.78-0.90)** | **0.83 (0.77-0.90)** | **0.83 (0.77-0.90)** | **0.84 (0.78-0.90)** |
|  | Qwen3-8B-Thinking | 0.74 (0.66-0.81) | 0.73 (0.66-0.81) | 0.74 (0.66-0.82) | 0.74 (0.67-0.81) |
| HPO+ICD+CoT | Llama-8B | 0.78 (0.72-0.87) | 0.80 (0.72-0.88) | 0.75 (0.69-0.84) | 0.78 (0.72-0.86) |
|  | Llama-70B | 0.8 (0.72-0.87) | 0.79 (0.72-0.86) | 0.77 (0.70-0.85) | 0.79 (0.72-0.86) |
|  | Qwen3-8B | 0.77 (0.69-0.84) | 0.76 (0.68-0.83) | 0.76 (0.67-0.83) | 0.77 (0.69-0.84) |
|  | Qwen3-8B-Thinking | 0.77 (0.69-0.85) | 0.76 (0.68-0.84) | 0.76 (0.67-0.84) | 0.77 (0.69-0.85) |

### Error Analysis: Why models make mistakes and how to reduce such mistakes

We performed a manual error analysis on 30 randomly selected test cases, reviewed by a board-certified clinical geneticist while initially blinded to the true labels. Review focused on our two best-performing fine-tune models, Qwen3-8B and Llama-3.1-8B, both configured with ICD features, CoT reasoning, and fine-tuned on summarized notes. The reviewer was asked to classify the prediction into one of the following categories: (1) Correct answer supported by valid CoT reasoning, (2) Correct answer but the CoT reasoning was wrong, (3) Incorrect answer but the CoT reasoning was informative, (4) Incorrect answer due to outdated gold standard and evolving guidelines, (5) Incorrect answer because of flawed CoT reasoning.

When computationally compared to existing gold-standard labels, 15 cases were correctly predicted by both models, 5 were correctly predicted by RareDAI-Qwen3-8B and another 5 by RareDAI-Llama-8B, and 5 were incorrectly predicted by both, yielding an apparent accuracy of 67% for each model. While this performance was lower than expected, expert review revealed that a substantial fraction of the apparent errors was attributable to limitations in the gold-standard annotations rather than model behavior.

For Qwen3-8B, the reviewer determined that 16 cases contained clinically valid reasoning with correct test recommendations, and 2 additional cases produced correct recommendations but incorrect reasoning. Among the remaining cases labeled as incorrect, 9 were found to conflict with the ground truth because the ground truth was based on outdated testing guidelines or evolving clinical practice rather than model error, while only 3 cases involved hallucinated reasoning leading to incorrect recommendations. Importantly, the clinician agreed with Qwen3-8B’s recommendations in 7 of the 9 cases affected by outdated gold standards. Accounting for these expert-validated cases, 23 of 30 responses demonstrated valid reasoning producing clinically appropriate recommendations (**Supplementary Figure 8**), corresponding to an adjusted accuracy of approximately 77%, substantially higher than the automated estimate.

A similar expert review of Llama-3.1-8B showed slightly stronger raw predictive performance, with 18 cases exhibiting correct predictions supported by valid reasoning and 1 additional case producing a correct recommendation but incorrect reasoning. Among the cases initially labeled as incorrect, clinicians agreed with the model’s recommendations in all 5 cases affected by outdated ground-truth annotations, while 4 cases were attributed to flawed chain-of-thought reasoning (**Supplementary Figure 8**). Interestingly there were two cases where the model provided the “correct” answer based on the gold-standard, but the reviewer agreed with the model’s CoT that no genetic testing was indicated by the available evidence. This variation would improve the actual accuracy after expert-reviews for the Llama model to 77%.

In addition, when evaluating models on raw clinical notes, errors stemmed from the model’s preference for genome sequencing in cases with broad symptoms, unclear family histories, and inconclusive prior tests. Misclassified patients with genome sequencing often had longer clinical notes (*mean value of* 4,496 ± 1,656 vs. 2,122 ± 384 tokens), more HPO (*mean value of* 58 ± 28 vs. 38 ± 17) and Phecodes (*mean value of* 8 ± 8 vs. 5 ± 4), introducing noise that reduced accuracy. Furthermore, some normal test results were mistakenly interpreted as justification for genome sequencing. For example, the model misclassified developmental and neurological tests as equivalent to gene panels, suggesting that a normal result from these tests indicates a need for genome sequencing—even though these tests are primarily psychological and imaging-based. In some cases, models exhibited hallucinated reasoning by incorrectly assuming that certain conditions represented genetic syndromes, resulting in unreliable chain-of-thought explanations.

Furthermore, the model analyzed a single (most recent before genetic testing) clinical note for each patient in isolation, to emulate the scenario where a diagnostic decision is made during a specific clinical encounter. However, clinicians may base their decisions on more than just the information in a single note. We recognize that important details could be buried in earlier clinical notes and not explicitly documented by genetic counselors or clinical geneticists during the encounter. We thus manually examined the notes for selected patients for whom RareDAI made a discordant recommendation and found that in some cases, there was no mention of family history or past diagnostic workups that could have informed the model’s decision. After tracing back through earlier notes, we discovered that key phenotypes from both the patient’s history and family history—potential indicators for genetic testing recommendations—were missing from the selected testing notes, which likely contributed to the discordant recommendations. This suggests that the model’s performance could be improved by incorporating longitudinal data rather than relying solely on the most recent clinical encounter.

### Comparison of Traditional Machine Learning and Interpretability Analysis

We compared RareDAI to Phen2Test^30^, a Random Forest–based model operating on structured phenotype and ICD-derived features. We retrained Phen2Test^30^, a Random Forest model developed by the Columbia team that leverages phenotypes, Phecodes, and clinical note counts to predict the appropriate genetic test (‘gene panel’ versus ‘WES/WGS’), using the same CHOP training cohort (N = 585) and evaluated it on an identical test set (N = 127). Phen2Test achieved an accuracy of 84% on the testing set. Across multiple configurations, RareDAI demonstrated comparable performance. When trained on summarized clinical notes augmented with ICD features and chain-of-thought (CoT) prompting, Llama-3.1-8B achieved the highest accuracy (86%), while Qwen-3-8B achieved 84%, matching the performance of Phen2Test (Table 3). In contrast, models trained on raw clinical notes exhibited reduced performance, likely due to increased input length and noise. Nevertheless, RareDAI remained competitive under these conditions: Llama-3.1-70B achieved 79% accuracy and Qwen-3-8B achieved 82% accuracy when incorporating HPO concepts, ICD features, and explicit reasoning (**Table 2**).

We further applied McNemar’s test to assess performance differences between RareDAI and Phen2Test (**Supplementary Figure 9**). While no statistically significant differences were observed between Phen2Test and the best-performing RareDAI configurations, the LLM-based approach provides complementary strengths beyond predictive accuracy, including flexible integration of unstructured clinical evidence and question-by-question, text-grounded reasoning.

To address interpretability, we analyzed Shapley value explanations from Phen2Test on testing samples and compared them qualitatively to RareDAI’s LLM-generated reasoning (**Supplementary Figure 10**). Shapley analysis identified high-importance structured features such as note counts, sensorineural hearing loss, age, autism, developmental delay, delayed milestones, and congenital anomalies of the skull and face, which are clinically plausible drivers of genetic testing decisions. These features frequently align with RareDAI’s explanations, which explicitly cite corresponding evidence extracted from clinical notes (e.g., descriptions of hearing impairment, neurodevelopmental delay, or craniofacial abnormalities) in the narrative format.

Finally, discrepancies between Shapley-based explanations and LLM reasoning arise naturally from a representational mismatch: Shapley explains model behavior within a closed, fixed feature space, whereas RareDAI reasons over open-ended natural language. By design, Phen2Test only sees 1,229 predefined structured variables including Phecodes, demographics, and note counts^30^. Meanwhile, our LLM-based framework produces question-by-question, ACMG-guided reasoning grounded directly in unstructured clinical notes, enabling clinicians to rapidly identify relevant supporting evidence for a testing decision. Because LLMs are trained to model the semantics of natural language, our approach is not restricted to a fixed feature set and can interpret phenotypes or Phecodes that extend beyond the predefined variables used by Phen2Test. These differences should not be interpreted as errors, but rather as complementary views of the decision process.

Overall, while Shapley values provide useful global and feature-level insights for Phen2Test, RareDAI offers instance-specific, clinically grounded explanations directly traceable to the patient’s narrative record. This distinction is particularly important for rare disease evaluation, where nuanced, heterogeneous, and incompletely structured phenotypic evidence frequently drives testing decisions.

## Discussion

The key objective of RareDAI was to utilize the chain-of-thought (CoT) generated by base LLMs to improve the fine-tuning process, addressing the shortage of medical experts for manual review. Given this lack of medical expertise, we used base LLMs to generate CoT answers for 585 training and 125 validation samples. We did not manually review all outputs for accuracy but randomly verified a few samples for coherence and data alignment. The Llama 3.1 70B-instruct and Qwen3-30B-A3B-Thinking-2507 models generated detailed observations, patient overviews, and reasonable label justifications. While unchecked synthetic explanations may introduce errors, our trials suggest that self-distillation is effective and yields promising results.

The results (**Tables 2 and 3**) indicate that, first, with CoT, the model can perform step-by-step analysis of patient symptoms and diagnoses, relying on the given test definitions, and then provides an accurate assessment. Second, it suggests that LLMs can generate reasoning-based answers. Given that the recommended genetic tests for the training samples are already known, it is reasonable to ask the model to reason and bias based on the prioritization of the true label over the alternative label. We instructed the model to leverage all relevant information that aligns with our predefined definitions and questions (**Supplementary Figure 2**). Despite not fine-tuning the models specifically for this generation task, the pretrained Llama 3.1 model produced a clear explanation due to its extensive pretraining and robust architecture.

As a complementary qualitative assessment, our clinical expert reviewed 30 randomly selected test cases to evaluate the plausibility and clinical appropriateness of the generated chain-of-thought reasoning. Overall, experts found that the models’ conclusions and reasoning aligned with clinical judgment in most cases. This review also revealed limitations of the original gold-standard annotations, which reflected clinical practice that has since evolved or was the product of clinical characteristics or family decision making not documented in the note.

When evaluated strictly against the original labels, both Llama-3.1-8B and Qwen3-8B achieved an accuracy of 67% on these samples. After expert adjudication, the proportion of cases judged to be clinically appropriate increased to 77%. These findings suggest that some apparent discrepancies between model predictions and reference labels may reflect annotation drift or incomplete documentation rather than clear reasoning errors, indicating that performance estimates based solely on static annotations may underestimate model behavior under current clinical standards.

Expert review further identified two cases in which genetic testing was not clinically indicated by the available evidence in the notes. Notably, Qwen3-8B explicitly recognized these cases, stating that no genetic test was warranted and only producing a recommendation when required to do so. The expert reviewer regarded this behavior as clinically appropriate, as it reflected an explicit acknowledgment of diagnostic uncertainty rather than forced binary decision-making. Across reviewed cases, clinicians also reported that Qwen3-8B produced more coherent, clinically grounded, and interpretable reasoning chains, underscoring the potential value of structured, text-grounded reasoning in clinical decision-support settings. However, to further improve this behavior, we recommend that future developers include samples with no recommended tests during training, thereby framing the task as a three-class classification problem.

Prior machine learning and neural NLP approaches have represented text using a wide range of techniques, including sparse feature representations (e.g., bag-of-words, word counts, TF–IDF), as well as dense word embeddings that predate modern transformer-based models^60,61^. These representations have been successfully used as inputs to many traditional and neural models, including non-transformer architectures.

Our comparison to conventional machine learning methods specifically refers to Phen2Test, which operates on predefined structured clinical features (e.g., phecodes and demographics variables) encoded using one-hot encodings, where each value is represented by a single “1” in a vector of zeros, limiting the richness of the data. While effective for leveraging structured data, this design requires information from clinical narratives to be abstracted into fixed categorical features and does not directly model contextual relationships within free-text notes.

In contrast, large language models build upon earlier embedding-based ideas by learning contextualized token representations that are dynamically conditioned on surrounding text and optimized end-to-end. Furthermore, Transformer-based models leverage attention mechanisms to establish relational importance between words in a sentence, enhancing the quality of generated answers^24^. Consequently, in this study, the decision between gene panel testing or genome sequencing for a patient is likely influenced by critical keywords within the context, such as whether the symptoms are specific to a single condition or associated with multiple conditions.

Excessive Phecodes do not necessarily improve LLM performance without strong reasoning analysis. Our study found that too many redundant Phecodes (more than 20) can inflate counts and degrade model performance, similar to overly long text inputs (typically greater 4500 tokens). We also evaluated how incorporating HPO terms affects model performance, since we have processed all clinical notes through cTAKES^62^ to extract structured HPO phenotypes. Models trained with phenotypic data showed mixed results. We found that that excessive phenotypic data—especially from automated NLP tools like cTAKES—introduces redundancies and negatively impacts smaller models like Llama-3.1-8B. Despite filtering, an average of 65 HPO terms per patient remained in our study, which is considerably higher than the <20 HPO terms typically recorded manually.

In contrast, HPO features were beneficial primarily when used with higher-capacity or reasoning-enabled models, such as Llama-3.1-70B or Qwen3-8B with thinking mode enabled. These models appear better able to filter and reason over noisy, redundant phenotypic inputs, whereas smaller models remain sensitive to feature inflation. Furthermore, as our external collaborator does not have cTakes-extracted HPO terms, we cannot evaluate whether it actually improves the performance. Future efforts should prioritize high-quality, manually curated phenotypic data (HPO terms) over NLP-generated data to enhance predictive accuracy while used as the prompts for LLMs.

We observe that clinical notes (even without additional ICD-10 or HPO) are often messy and unstructured, and lengthy. It is well known that LLM often degrades the performance when inputs contain extraneous texts such as administrative artifacts, templates, or duplicated documentation^63,64^. We hypothesized shortening notes to be more succinct while retaining quality information would improve performance and designed a prompt directing the LLM to summarize clinical notes based on key categories (**Supplementary Figure 4**). Our analysis (**Tables 2 and 3**) shows that LLMs perform better with summarized clinical notes, achieving the highest performance (achieving 86% accuracy and 85% F1-score for RareDAI-Llama 8B) across all configurations. We further applied McNemar’s test to assess whether pairwise differences in model predictions reflect statistically significant improvements rather than random variation on the same test set (**Supplementary Figure 9**). McNemar’s test reveals no statistically significant differences among models trained on the same input representation, including comparisons between full-note models and between summarized-note models (most p-values > 0.1; **Supplementary Figure 9**). In contrast, statistically significant differences consistently emerge when comparing models trained on summarized notes against those trained on full notes, indicating that input representation—rather than model architecture alone—is the primary driver of meaningful performance differences. Shorter, structured inputs reduce noise and redundant tokens, improving the model’s reasoning and pattern recognition. Since LLMs rely on contextual token prediction, minimizing irrelevant information enables smoother, more accurate inference. Although performance was lower on raw notes, studying their use as input offers deeper insights into real-world challenges. Overall, these results highlight the value of strategic data preprocessing, showing that careful summarization significantly enhances model efficiency and predictive performance when analyzing complex medical records.

RareDAI was trained and evaluated on clinical notes with patient-level demographic diversity, as shown in **Supplementary Figure 1**. While the dataset includes variation in sex, race, ethnicity, and age, the distributions are imbalanced—for example, White and non-Hispanic patients are overrepresented relative to other subgroups. Since RareDAI takes raw notes as input and produces genetic testing recommendations as output, such imbalances raise the possibility of subgroup-specific bias, where recommendations may generalize less effectively to underrepresented populations. To mitigate this risk, future work will stratify RareDAI’s performance by demographic categories and apply fairness-aware modeling approaches. We also emphasize that RareDAI is intended as a decision-support tool, not a replacement for clinical judgment, and careful monitoring of subgroup outcomes will be essential to ensure equitable and responsible use in genomic medicine.

As with any clinical decision support system, incorrect recommendations from RareDAI may carry potential downstream harm if used without appropriate safeguards. In the context of genetic testing, a false recommendation for genome sequencing may lead to unnecessary cost, increased turnaround time, and patient anxiety, whereas an incorrect recommendation for a more limited gene panel could delay diagnosis or necessitate repeat testing. Importantly, RareDAI is designed to function as a decision support tool rather than an autonomous decision-maker, and its outputs are intended to augment—rather than replace—clinician judgment. Several design choices mitigate these risks. First, RareDAI provides evidence-grounded, structured explanations that allow clinicians to inspect the underlying rationale and identify potential errors or mismatches with clinical context. Our human evaluation suggests that even in some misclassified cases, the model highlights clinically relevant evidence that can still inform expert decision-making. Second, the system is optimized to support second-opinion scenarios or triage rather than final test ordering, reducing the likelihood that a single incorrect recommendation directly translates into clinical action. Third, the use of question-specific reasoning enables clinicians to recognize which aspects of the recommendation (e.g., phenotype complexity, diagnostic uncertainty, cost considerations) are driving the model’s output and to override individual components when appropriate. Future work will further reduce potential harm by incorporating uncertainty-aware outputs, stricter evidence-presence constraints for specific questions, and workflow-level safeguards such as requiring explicit clinician confirmation before action. Together, these measures position RareDAI as a transparent and clinically responsible support tool, while acknowledging the importance of human oversight in genetic testing decisions.

From a clinical perspective and qualitative expert review, questions related to congenital/developmental abnormalities (Question 1), multigenic or complex phenotypes (Question 2), and prior inconclusive testing (Question 5) were the most influential for accurate genetic testing recommendations, as they directly align with ACMG criteria for escalation to exome or genome sequencing. Based on qualitative inspection during error analysis and expert review, hallucinations were more likely to arise in questions that require strong assumptions about phenotype specificity or external constraints, particularly Question 3 (distinctive features suggesting a specific condition) and Question 7 (cost or accessibility considerations). These questions depend on implicit clinical judgment or contextual information that is often incompletely documented in clinical notes, making them more susceptible to over-interpretation by the LLM. From manual review, sometimes, models incorrectly assumed certain conditions are genetic or development syndromes (e.g., Qwen3 misclassified hearing loss as a developmental syndrome). In contrast, questions grounded in clearly documented clinical facts (e.g., congenital anomalies, prior testing, or urgent care settings) were generally more stable. However, such errors can often be corrected through clinician judgment at the point of use, while preserving the validity of the overall reasoning approach.

Despite the best prediction accuracy of 86%, RareDAI demonstrates strong precision where it matters most. Among the 127 patients evaluated, we have information on a subset of 21 (16.5%) who received positive genetic diagnosis (we do not have information on others), including 9 via gene panel and 12 via WES/WGS. Remarkably, RareDAI correctly recommended the test that led to diagnosis for all 21 patients, achieving a 100% alignment. While the sample size is modest, this analysis suggests that RareDAI captures core decision-making patterns used by clinical geneticists and is reliable when the stakes are highest, highlighting its strong potential to support and scale expert-level clinical reasoning. This finding reinforces RareDAI’s value as a triage or decision support tool, as the model consistently prioritizes the right test for the patients who ultimately benefit from it.

The potential clinical utility of RareDAI lies in its role as a decision-support tool that can assist clinicians by providing relevant clinical evidence and guideline-informed reasoning to support test selection. In current clinical workflows, test selection often depends on access to specialized expertise, which can delay diagnosis and strain genetics services. RareDAI could potentially augment this process by providing expert-level reasoning into a scalable model that interprets diverse clinical inputs—structured and unstructured—and recommends the most appropriate test pathway based on established standards such as the ACMG guidelines.

In the future, such recommendation technology could be incorporated directly into a clinical workflow: clinicians would input a patient’s clinical note, and the system would generate a structured chain-of-thought explanation along with a recommended test category (e.g., gene panel vs. genome sequencing). The clinicians would then review this reasoning alongside their own assessment, using it as an additional layer of decision support rather than a replacement for clinical judgment. This framework not only clarifies why one test may be more appropriate than another—helping navigate critical trade-offs such as when to escalate from a panel to WES/WGS—but also provides justification for resource allocation and insurance approval. In environments where time, expertise, or access to genetics specialists is limited, RareDAI can act as a scalable second-opinion tool, potentially reducing unnecessary broad testing, optimizing resource use, and ultimately lowering healthcare costs.

As mentioned in our error analysis above, training on a single clinical note per patient may not capture the full complexity of a case, which subsequently affects recommendation accuracy. In this study, we deliberately selected Progress Notes and in rare cases History & Physical (H&P) Notes, and further prioritized most recent note prior to the genetic testing recommendation (usually the highest word counts) under the assumption that these note types at CHOP routinely carry forward salient historical information from earlier encounters. Despite these design choices, our manual review of misclassified cases indicates that important longitudinal context—such as symptom progression, earlier differential diagnoses, or prior testing considerations—can be fragmented across encounters and absent from the selected note. This is perhaps the most significant challenge and may explain why the model achieves only 70-86% across all metrics. Future work could reduce this source of error by more selectively filtering notes by clinical department (e.g., genetics-specific encounters) or by aggregating non-redundant information from multiple time-stamped notes using temporal pooling or hierarchical summarization strategies prior to inference with RareDAI.

Filtering irrelevant information from clinical notes remains difficult, yet this irrelevant information may introduce noise for LLMs and may mislead LLMs when making the correct predictions. The challenge is compounded by memory constraints of processing long notes. To address these challenges, we utilized a data summarization strategy to generate summarized versions of clinical notes and reduce text size, before fine-tuning and making inference. Our preliminary results show that this strategy improves performance, and we imagine that improved summarization approaches in the future can help RareDAI to further improve performance. Moreover, text-based assessments in RareDAI (even by integrating structured information) also have limitations, as descriptions of physical features may be insufficient for accurate recommendations. Integrating multimodal data, such as images or videos, could enhance performance in future studies. Moreover, some factors influencing the choice of testing strategy—such as patient preferences—may not be documented in the clinical notes. Finally, the model was fine-tuned on clinical notes from a single institution (CHOP), potentially restricting generalizability. Notably, the performance of our model on an independent dataset from Columbia is lower than the CHOP data. Since documentation styles vary across healthcare systems, institution-specific models may be necessary for optimal accuracy. To address this limitation, we have made our computational procedures and open-source software code publicly available at GitHub, so that users can reproduce our self-distillation fine-tuning strategy on local datasets to ensure reliable performance in real-world applications. Genetic testing methods, including cost and turn around time, are constantly changing with the rapid development of sequencing technology, so this may lead to different optimal choices over time. Finally, RareDAI used clinical assessments generated by genetic counselors and clinical geneticist for training and at inference time. If RareDAI were to be used more broadly as a clinical decision support tool, lack of expert documentation would be a key barrier.

Although few-shot in-context prompting has demonstrated benefits in prior clinical NLP studies^65–68^, we did not adopt this strategy in the current work due to several task-specific considerations. Our experiments and prior study^64^ indicate that LLM performance degrades substantially when operating on very long clinical notes, even when the nominal context window is sufficient. Applying few-shot prompting in this setting would require including at least two full examples—one for genome sequencing and one for gene panel to avoid label bias—each consisting of a complete clinical note, multi-step reasoning, and final recommendation, which can span several thousand tokens per example. This would substantially increase prompt length and may further degrade performance, as earlier tokens receive progressively weaker effective attention as context grows, especially for lightweight models (8B). In addition, based on preliminary observations and prior experience, embedding full clinical examples in the prompt can increase the risk that models inadvertently reuse or blend example-specific details into the target output, potentially introducing spurious or hallucinated information, while constructing expert-validated exemplar explanations at scale remains challenging. Instead, we adopted a structured, question-guided prompting strategy that provides explicit definitions of gene panel and genome sequencing, together with clinically motivated guiding questions (e.g., ACMG-aligned criteria), to ground model reasoning without relying on full example cases. This approach offers a more scalable and controlled alternative for long-form clinical text, while future work may explore lightweight or hybrid few-shot strategies that mitigate these challenges. Regardless, we acknowledge the value of few-shot prompting in clinical NLP, and therefore evaluated a simple two-example few-shot setting using ChatGPT@CHOP (**Supplementary Figure 11**) on the same 30 manually curated notes used in our RareDAI evaluation (but without expert verification). Adding one exemplar for gene panel and one for genome sequencing improved gold-standard accuracy by ∼10% compared to the zero-shot setting. When evaluated against the clinically adjusted labels, ChatGPT@CHOP achieved 83% accuracy, outperforming our best fine-tuned open models. These results indicate that few-shot prompting can remain effective even under long-context clinical inputs, at least for a highly capable model. However, this performance should be interpreted in the context of substantial system-level differences. While the underlying architecture and parameter count of ChatGPT@CHOP (version 5.0) are not publicly disclosed, it likely benefits from substantially larger model capacity than 8B-scale open models (hundreds of billions to trillions), stronger proprietary pretraining, and additional long-context robustness from large-scale instruction tuning and optimization. As a result, the degree to which few-shot prompting helps in ChatGPT@CHOP may not directly transfer to lightweight public models fine-tuned for this task, where adding full exemplars can meaningfully increase prompt length and potentially interfere with evidence grounding (e.g., by encouraging exemplar-style reasoning or blending exemplar-specific details into the target output). Moreover, HIPPA-compliant ChatGPT is available only within certain institutional environments, limiting reproducibility and broad deployment across healthcare systems. In contrast, our work prioritizes accessibility and deployability by using fully public models that can be deployed on-premise and applied to institutional clinical notes without exposing PHI. Despite operating under a stricter exemplar-free prompting setup, our fine-tuned models remain competitive and produce clinically grounded reasoning, supporting task-specific fine-tuning as a practical and scalable alternative when proprietary systems or institution-specific access to ChatGPT@CHOP are unavailable.

In this work, we used a fixed set of seven ACMG-guided chain-of-thought questions and did not explore automated prompt optimization or question discovery. Techniques such as gradient-based prompt optimization (e.g., TextGrad^69^) could be used in future work to refine or adapt the CoT prompts and to study how prompt structure influences both accuracy and reasoning quality. In addition, we did not explicitly investigate prompt-masking or prompt-level loss weighting during training; while the CoT prompts were included in the training input, we did not isolate or reweight their contribution to the loss. Exploring prompt-aware training strategies and evaluating model robustness to alternative or newly generated questions are important directions for assessing generalization beyond the predefined CoT framework.

## Methods

### Ethics approval and consent to participate

This study was approved by the Institutional Review Board of the Children’s Hospital of Philadelphia (IRB #18-015712). All research procedures complied with institutional ethical guidelines and relevant regulations governing human subjects research. A waiver of informed consent and assent was approved under the study protocol because the research was retrospective and posed minimal risk to participants. All computational analyses were conducted within an institutionally approved secure computing environment in accordance with data governance and privacy guidelines.

### Study Cohort

Leveraging the electronic health records available, clinical notes, structured phenotypes, and Phecodes (expert-defined groups of International Classification of Diseases, 10th revision (ICD-10)) were incorporated into both the training and evaluation processes. The data for this study at CHOP was accessed through the Arcus platform, which seamlessly links biological, clinical, research, and environmental data^42^. Genetic counselors and clinical geneticists examined patients and made recommendations for patients to undergo either gene panel testing, exome/genome sequencing, or both.

The original cohort for training and validation consisted of 997 patients at CHOP, but after applying exclusion criteria, we reduced this to 837 patients (shown in **Supplementary Figure 1**). To ensure unbiased model training and avoid incorrect pattern learning, we excluded notes mentioning specific genetic tests and filtered for high-quality documentation like “Progress Notes” and “H&P Notes” (History & Physical), which provide rich clinical detail. Specifically, we removed any notes containing the keywords “gene panel”, “wes”, “wgs”, “exome sequencing”, or “genome sequencing” (case-insensitive). This exclusion step was implemented to prevent label leakage and ensure that the model did not “cheat” by learning direct test names rather than reasoning from patient context. To confirm the effectiveness of this approach, we manually spot-checked a random subset of excluded and retained notes, which verified that the filtering successfully removed direct mentions while preserving clinical content. We further refined the dataset by including only notes with 600–5000 words and using a consistent starting pattern for untagged notes from CHOP to ensure relevance and completeness. Through observations, notes under 600 words often lack meaningful information, typically containing discussions between the patient’s family and medical staff regarding appointments, follow-ups, medications, or treatments, which are less relevant for genetic test determination. We set the upper limit at 5000 words due to memory constraints during model training. Although we experimented with notes up to 7000 words, models struggled to train with any collections with many notes exceeding 5000 words. For each patient, we selected only the notes with the highest word count that were filed before the date of genetic recommendations by IMGC specialists. In cases where a patient was recommended for two tests (N_train_ = 28, N_val_ = 8), we chose the ultimate recommendation, as we believe that the specialists had sufficient time during the gap period to assess the patients more thoroughly and make informed decisions. Hence, the ultimate test recommendation could reflect the most comprehensive overview of the patient’s clinical presentation, prior diagnostic outcomes, and evolving medical understanding. This approach allows us to anchor our model training on decisions that are likely informed by the most complete and mature clinical reasoning available within each case.

Among them, 510 underwent genetic testing with exome/genome sequencing and 327 underwent genetic tests with gene panels. We randomly divided the cohort into training, validation, and test sets in a 6:2:2 ratio (585:125:127). **Supplementary Figure 1** shows histograms for word counts and token counts across the three sets. Details on how we curated and processed data are illustrated in the Supplementary Note. We further converted ICD-10 codes into Phecodes^43,44^. Classification models for diverse clinical genetic applications have been built using Phecodes^37^, hence, we consider them as a valuable input option. On average, each patient was assigned 9 Phecodes, allowing for efficient model training without memory constraints. Similarly, applying the same approach to phenotypes showed that this group still had a high phenotype burden, with an average of 65 phenotypes per patient. When patients have no recorded ICD-10 diagnosis or phenotypes, the phrase “Please refer to the note for more detailed information” is used.

Clinical notes are inherently noisy and unstructured, making it difficult to standardize their format. Variations in note structure often arise due to differences in authors, the use of different templates, and the underlying EHR system. One of the key objectives of this project is to allow LLMs to learn the complexities of these unstructured clinical notes and provide explainable recommendations. By doing so, future users can easily input raw data without undergoing complicated preprocessing steps.

Our fine-tuned model was further evaluated on an independent cohort from Columbia University Irving Medical Center (CUIMC). The dataset includes Phecodes and clinical notes from 766 patients—421 of whom underwent exome/genome sequencing and 345 of them received gene panels. Each patient has 8 Phecodes on average, with summarized clinical notes containing 403 words on average.

### Pretrained Large Language Models – Llama 3.1

Over the past two years, LLMs have transformed the deep learning landscape, with Meta’s Llama models among the most powerful open-source options^26^. The mid-2024 Llama 3.1 family of models was released in three variants: 8B, 70B, and 405B parameters each with instruction fine-tuned variants. Throughout this manuscript, we use only the instruction fine-tuned variants but refer to them only by their base model identifier. Although the training dataset is not publicly available, Llama models were pretrained on extensive text corpora, including English Common Crawl, C4, GitHub, public datasets, and some medical texts^26,45,46^.

While the 405B model excels in benchmarks, its high memory demands make fine-tuning difficult. The 8B and 70B models offer a balance between performance and efficiency. One of our key objectives is to offer the community a feasible strategy to fine-tune open-sourced LLMs capable of providing reliable genetic testing recommendations so that the users can replicate the whole process with their in-house data. To optimize fine-tuning, we apply techniques such as LoRA^47^ (Low-Rank Adaptation) in 8-bit precision, QLoRA^48^ (Quantized Low-Rank Adaptation) in 4-bit precision, and PEFT^49^ (Parameter-Efficient Fine-Tuning), allowing us to fine-tune models effectively with our existing resources. Higher precision improves performance but demands more resources. In both LoRA and QLoRA configurations, we only fine-tuned the adapter layers with the rank of 128 while all layers were fine-tuned in the full-finetuning models. Regarding computational costs, fine-tuning the Llama 8B model in 16-bit precision required approximately 6 hours on four A100-40GB GPUs, whereas fine-tuning the Qwen3-8B (full-parameters) or Llama 70B model with QLoRA (4-bit) and PEFT required eight H100 GPUs and ∼14 hours for the same dataset (585 training, 125 validation notes). While higher precision generally improves performance, it comes at a significant increase in resource demand. These comparisons highlight the trade-off between efficiency and accuracy: smaller models are more cost-effective, whereas larger models achieve stronger reasoning but require substantially greater computational resources.

### Pretrained Large Language Models – Qwen 3

In addition to Llama, we employed models from the Qwen 3 family^50^ developed by Alibaba, which have demonstrated strong instruction-following and reasoning capabilities. Specifically, we fine-tuned the Qwen3-8B dense model, which offers a favorable balance between performance and computational efficiency, making it suitable for in-house training. Separately, we used the Qwen3-30B-A3B-Instruct-2507 model, a mixture-of-experts architecture with approximately 30B total and 3B active parameters per token, to generate chain-of-thought (CoT) rationales; this model was not fine-tuned and was used solely for inference. Both models were pretrained on large-scale multilingual corpora spanning web text, code, and reasoning-focused data^50^, and their complementary roles allowed us to leverage efficient training while benefiting from stronger reasoning during CoT generation.

### Optional Clinical Note Summarization Step

In this study, we evaluated the performance of RareDAI on both raw and summarized clinical notes. To transform raw, noisy, and often lengthy clinical notes into structured summaries containing the essential information required for genetic testing recommendations, we applied an AI-based summarization step prior to model training and evaluation. This process was guided by a structured prompt consisting of predefined, clinically motivated questions (**Supplementary Figure 4**), which encourages the model to focus on specific, decision-relevant portions of the clinical notes rather than summarizing all content indiscriminately. By constraining the extraction to relevant phenotypic evidence, diagnostic context, and testing-related considerations, this approach may help limit the inclusion of irrelevant details and reduce the likelihood of introducing unsupported information into the summary. When referring to “summarized notes”, we mean notes that are intentionally transformed into a structured, concise representation organized around predefined clinical questions, while preserving all information required for genetic testing recommendations.

This structured summarization facilitates downstream modeling and improves robustness across institutions. We applied the same summarization procedure when validating RareDAI on the external CUIMC cohort, which helps reduce potential privacy risks, minimize inadvertent data leakage, and promote a more consistent, institution-agnostic input representation. In this study, we used Llama-3.1-70B model to generate summarized notes for fine-tuned Llama family while Qwen3-30B-A3B-Thinking-2507 was used for fine-tuned RareDAI-Qwen3 variant.

### Chain-of-Thought (CoT)

One of the effective techniques for enhancing the quality of LLM responses is the use of chain-of-thought (CoT) reasoning^38–41^. Given that models rely on next-token prediction, the preceding words heavily influence the generation of subsequent tokens. CoT helps LLMs emulate human-like brainstorming processes before reaching a conclusion. Previous studies have demonstrated that when given specific output, like a genetic testing decisions in our cases, base LLMs can effectively express the reasoning for why the output was generated^40,41^. Therefore, in this study, we leveraged the Llama-3.1-70B model and Qwen3-30B-A3B-Thinking-2507 to generate CoT-style responses to enhance our training of the model.

To improve model reasoning, we prompted the LLMs to explain why the correct genetic test (true label) was preferable given the patient input, using our seven questions (**Figure 1**) inspired by ACMG guidelines and previous studies^1,51^. This structured approach helps the model extract key information and generate well-structured chain-of-thought (CoT) responses. These questions were later used in both fine-tuning and inference to teach the model how to derive accurate answers. The CoT responses generated were integrated as part of the desired output into fine-tuning, allowing the model to learn logical explanation construction before making decisions. This fine-tuning technique is known as self-distillation fine-tuning (SDFT)^52^ (**Figure 1**). **Supplementary Figure 2** illustrates the prompt template used to generate CoT explanations for the primary fine-tuning process. The features (predictors in a machine-learning model) included in the fine-tuning process, such as Phecodes, must align precisely with those used during the CoT generation. To preserve semantic information, we provide Phecodes in text form rather than using their corresponding IDs.

### Fine-tuning Template

Fine-tuning LLMs can enhance performance, especially in domain-specific settings such as clinical genetics. A key impetus for fine-tuning is to imbue base models with specialized domain-specific knowledge or skills such clinical notes interpretation or genetic diagnosis. Since base LLMs are pretrained on general corpora and not specifically designed for molecular genetic testing, we included definitions of key terms like gene panel and genome sequencing from medical sources^14^. The user prompt contains seven guiding questions along with relevant inputs—Phecodes and clinical notes—used consistently across training, validation, and testing. The assistant prompt, included only during training, helps the model learn how to approach the main question using the provided inputs. In this phase, the LLM was fined-tuned to generate responses, incorporating a CoT explanation when available. **Supplementary Figure 3** illustrates the template with our prepared questions and sample inputs.

## Data availability

The raw clinical data used to train the model cannot be shared publicly to protect patient privacy at the Children’s Hospital of Philadelphia. To support reproducibility, we provide the full implementation and processing pipeline on GitHub, and external users may replicate the workflow using their own institutionally approved clinical notes and datasets.

## Code availability

The instructions and codes in this study are available on GitHub at https://github.com/WGLab/RareDAI. The repository provides a configuration file for users to setup the virtual environment with the required Python packages installed to run the scripts.

## Acknowledgements

We used OpenAI’s ChatGPT@CHOP (GPT-5, July 2024 version) to improve the readability and flow of the manuscript. The original ideas, content, and analyses were entirely developed by the authors, and no scientific or conceptual discussions were conducted with the model. The exact prompt used was: “Polish the following paragraph for clarity and conciseness while preserving the original meaning.” This tool was used selectively to refine language in the Introduction, Methods, Results, and Discussion sections. This study was supported by NIH grants HG012655, HG013031, HD111688 and the CHOP Research Institute.

## Author Contributions

The author contributions in this study are outlined as follows: Q.M.N led the project’s conception and defined the research aims, data collection and curation, performed label annotation and validation, handled data preprocessing and analysis, developed and tested the models, explored the outcomes, created data visualizations, and drafted the initial manuscript. K.S., S.E.S, and I.M.C provided the list of patients with label annotations. K.W., W.C., C.W., C.L., D.W., I.M.C and C.N.T. supported methodology development and contributed to manuscript revisions. I.M.C, a board-certified clinical geneticist, manually verified the CoT from 30 testing samples generated by fine-tuned Llama-3.1-8B and Qwen3-8B. F.C., G.Z., P.A., and C.L. assisted with validation on the Columbia dataset. K.W supervised the project, shaped the research concept and scope, managed project administration, oversaw algorithm development and validation, secured funding, and revised the manuscript. All authors reviewed and approved the final manuscript.

