## Supplementary Figures for "Interpretable Fine-tuned Large Language Models Facilitate Making Genetic Test Decisions for Rare Diseases"

### Supplementary Note

#### Model diagnostics: feature importance, and quantization levels in RareDAI

In addition to clinical notes, RareDAI also incorporates structured phenotype information, such as Phecodes (converted from ICD-10 codes). We next examined how structured information contributes to model performance. Without CoT, incorporating the ICD-10 diagnosis did not improve model performance, as evidenced by relatively minor change across all metrics in Note/ICD models compared to 'Note Only' (Standard Supervised Fine-tuning method), in both Llama models. However, if we integrate CoT then it can enhance the models (CoT/ICD), making them the top performers. Moreover, the addition of phenotypes alone with CoT does not significantly enhance model performance. In fact, the 8B model experiences a notable decrease in scores, with reductions of 3-6% across all metrics. However, it is important to highlight that the CoT/P/ICD 70B 4-bit model achieves the highest performance, reaching 79% in accuracy, 78% in precision and 77% in recall and F1-score among all models in this study when analyzing the long unstructured notes. This suggests that this variant excels in reasoning analysis and accurately identifying samples recommended for genetic tests.

Furthermore, we evaluated how quantization levels impact model performance. Quantization is used to reduce the memory and computational requirements of LLMs by converting high-precision numerical values (e.g., 32-bit floating-point numbers) into lower-precision representations (e.g., 8-bit integers). This process reduces hardware requirements and speeds up inference but may compromise accuracy. **Supplementary Figure 12** shows that the 8B model with LoRA 8-bit (roughly 70% across all metrics) underperformed the half precision (16 bit) fine-tuned version (averagely 74% across all metrics). We also compared the base 70B model in both half precision and 4-bit quantized versions. The base 4-bit quantized 70B model performed poorly compared to the 16-bit base 70B model. This suggests that lower-bit quantized models may lack the capacity to provide meaningful explanations and accurate assessments for genetic recommendations. Therefore, further fine-tuning of the 70B model with LoRA 8-bit or even half precision optimization, along with phenotypes, phecodes, and CoT, remains a promising path for improvement if the users' computing resources allow.

### Supplementary Figures

**Supplementary Figure 1**: Demographic (Sex, Race, Ethnicity, Age), Word and Token Length Distributions Across CHOP Train, Validation and Test datasets. Note that: the generated CoT and labels are incorporated in Train and Validation sets while Test set includes the input only.

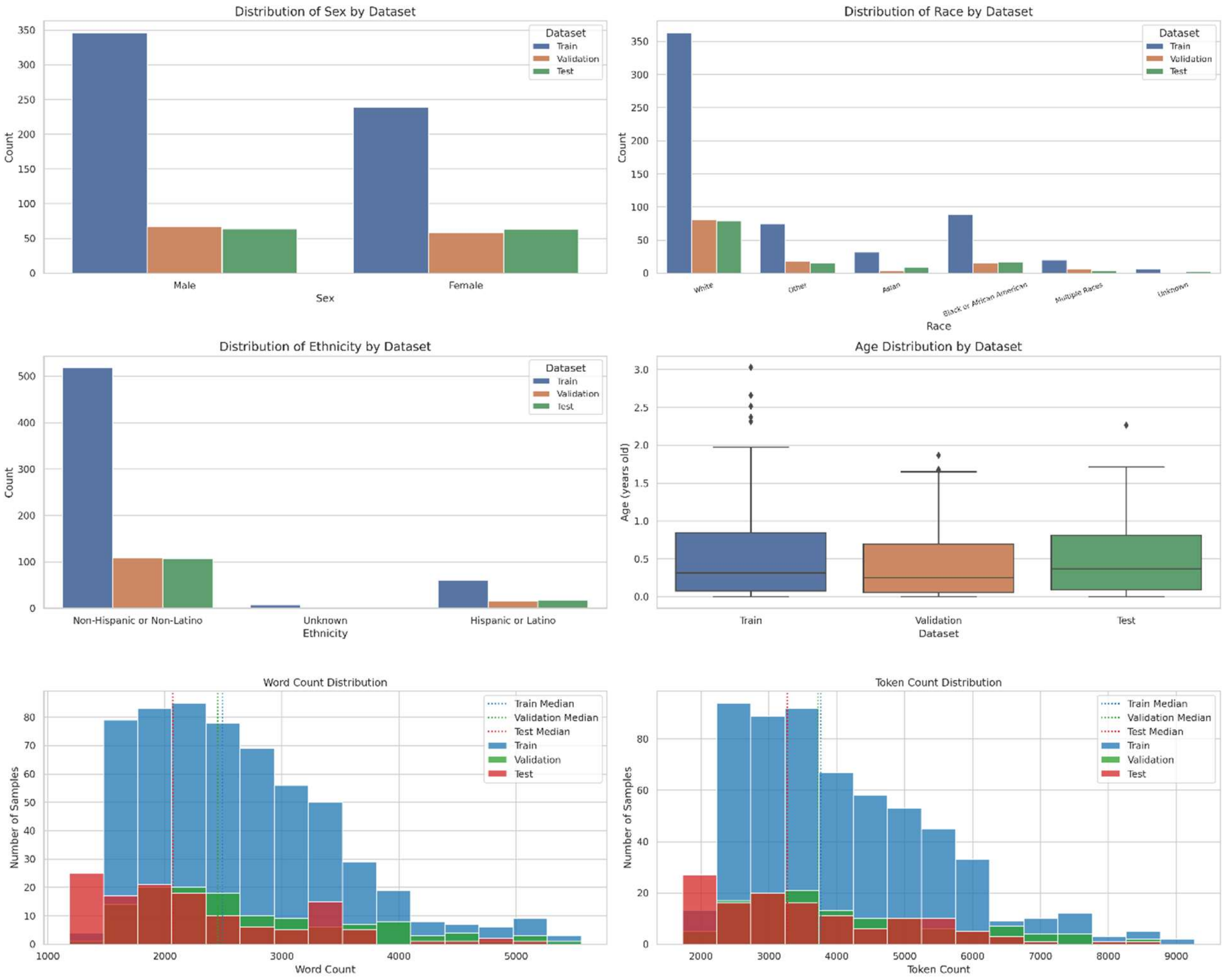

**Supplementary Figure 2**: This template is used to GENERATE chain-of-thought responses from the base Llama 3.1 70B-Instruct model and Qwen3-30B-A3B-Instruct-2507, serving as a preliminary step before the actual fine-tuning process. We want the model to be purposely reasoning in favor of the true genetic test over the false label. The explanations generated at this stage are then integrated into the main fine-tuning phase.

```
<|begin_of_text|>
<|start_header_id|>system<|end_header_id|>
You are a genetic counselor adhering to the standards and guidelines set by the American College
of Medical Genetics (ACMG). Given the following definitions, answer the question concisely and
don't make up any random answer.
gene panel: look for variants in more than one gene. This type of test is often used to pinpoint
a diagnosis when a person has symptoms that may fit a wide array of conditions, or when the
suspected condition can be caused by variants in many genes. Gene panel is suitable for any
patients with distinctive clinical features, family history of a specific disorder, or indicative
biochemistry, X ray, or complementary assays in a fast manner and less expensive than exome
sequencing.
genome sequencing: analyze the bulk of an individual's DNA to find genetic variations. Whole
exome or whole genome sequencing is typically used when single gene or panel testing has not
provided a diagnosis, or when the suspected condition or genetic cause is unclear. Genome
sequencing is often more accurate and applicable for patients with multiple nonspecific concerns
but takes longer to be done.

<|eot_id|>
<|start_header_id|>user<|end_header_id|>
Explain why we recommend {TRUE_LABEL} as the best choice for the patient described in the
following clinical note. Build a detailed, expert, and logical step-by-step reasoning explanation
for why {TRUE_LABEL} is strongly preferable to {FALSE_LABEL} in this case to convince our
physicians and patients from all the given phenotypes and clinical note. You should rely on the
given questions to build your logical answer.
1. Is the patient presenting with congenital abnormalities or developmental disorders? According
to ACMG guidelines, exome or genome sequencing should be considered as a first or second-tier
test for these conditions to provide a comprehensive diagnostic approach.
2. Does the patient's condition or suspected genetic disorder involve multiple genes or a complex
phenotype that cannot be confidently explained by a single-gene or targeted panel approach? If
yes, ACMG guidelines support the use of exome or genome sequencing for broader evaluation and
potential reanalysis over time.
3. Does the patient have distinctive clinical features, biochemical findings, or imaging results
that suggest a particular genetic condition or set of conditions? If yes, a targeted gene panel
may be the most efficient approach. If no, exome or genome sequencing might be more appropriate
to uncover a broader range of potential genetic causes.
4. Does the patient have a high likelihood of a specific genetic disorder based on family
history? If yes, ACMG guidelines recommend starting with targeted testing or a gene panel to
efficiently identify causative variants.
5. Has the patient undergone prior genetic testing or any diagnostic tools, and were the results
inconclusive or insufficient for diagnosis? If yes, exome or genome sequencing should be
considered to provide a broader diagnostic perspective. If no, a gene panel may be the first
step, especially if the phenotype suggests a specific set of conditions.
6. Is the patient in an urgent care setting, such as the NICU or ICU, requiring rapid results for
clinical management? If yes, rapid exome or genome sequencing is preferred for its ability to
quickly evaluate most protein-coding genes at once.
7. Are there cost or accessibility concerns that might limit the use of exome or genome
sequencing as a first-tier test? If yes, ACMG guidelines suggest that a targeted approach, such
as a gene panel, may be a pragmatic starting point while considering sequencing as a follow-up.
Input:
|==|ICD-10 Diagnosis|==|
Phecode X | Phecode Y | Phecode Z | Phecode W
|==|Note|==|
{clinical note here}

<|eot_id|>
<|start_header_id|>assistant<|end_header_id|>
|==|Explanation|==|
{CoT}
```

**Supplementary Figure 3**: This template is designed to FINE-TUNE Llama models for a specific task using CoT explanations generated from the previous stage. The key distinction between this template and the CoT Generation template lies in the user prompt. In the prior stage, since the training and validation sets already have known labels, we leveraged the LLM's reasoning capabilities to generate analyses rather than manually annotating them. These analyses were then used to train the model to answer the primary question: "What genetic testing do you recommend for the patient?" Fine-tuning ensures that the model mimics these analyses while also aligning human preferences in response style when it faces the new query.

```
<|begin_of_text|>
<|start_header_id|>system<|end_header_id|>
You are a genetic counselor adhering to the standards and guidelines set by the American College
of Medical Genetics (ACMG). Given the following definitions, answer the question concisely and
don't make up any random answer.
gene panel: look for variants in more than one gene. This type of test is often used to pinpoint
a diagnosis when a person has symptoms that may fit a wide array of conditions, or when the
suspected condition can be caused by variants in many genes. Gene panel is suitable for any
patients with distinctive clinical features, family history of a specific disorder, or indicative
biochemistry, X ray, or complementary assays in a fast manner and less expensive than exome
sequencing.
genome sequencing: analyze the bulk of an individual's DNA to find genetic variations. Whole
exome or whole genome sequencing is typically used when single gene or panel testing has not
provided a diagnosis, or when the suspected condition or genetic cause is unclear. Genome
sequencing is often more accurate and applicable for patients with multiple nonspecific concerns
but takes longer to be done.

<|eot_id|>
<|start_header_id|>user<|end_header_id|>
What genetic testing do you recommend to the patient in the following input? Please give a
detailed explanation and return 'gene panel' or 'genome sequencing' as the final response. You
should rely on the given questions below to build your logical answer.
1. Is the patient presenting with congenital abnormalities or developmental disorders? According
to ACMG guidelines, exome or genome sequencing should be considered as a first or second-tier
test for these conditions to provide a comprehensive diagnostic approach.
2. Does the patient's condition or suspected genetic disorder involve multiple genes or a complex
phenotype that cannot be confidently explained by a single-gene or targeted panel approach? If
yes, ACMG guidelines support the use of exome or genome sequencing for broader evaluation and
potential reanalysis over time.
3. Does the patient have distinctive clinical features, biochemical findings, or imaging results
that suggest a particular genetic condition or set of conditions? If yes, a targeted gene panel
may be the most efficient approach. If no, exome or genome sequencing might be more appropriate
to uncover a broader range of potential genetic causes.
4. Does the patient have a high likelihood of a specific genetic disorder based on family
history? If yes, ACMG guidelines recommend starting with targeted testing or a gene panel to
efficiently identify causative variants.
5. Has the patient undergone prior genetic testing or any diagnostic tools, and were the results
inconclusive or insufficient for diagnosis? If yes, exome or genome sequencing should be
considered to provide a broader diagnostic perspective. If no, a gene panel may be the first
step, especially if the phenotype suggests a specific set of conditions.
6. Is the patient in an urgent care setting, such as the NICU or ICU, requiring rapid results for
clinical management? If yes, rapid exome or genome sequencing is preferred for its ability to
quickly evaluate most protein-coding genes at once.
7. Are there cost or accessibility concerns that might limit the use of exome or genome
sequencing as a first-tier test? If yes, ACMG guidelines suggest that a targeted approach, such
as a gene panel, may be a pragmatic starting point while considering sequencing as a follow-up.
Input:
|==|ICD-10 Diagnosis|==|
Phecode X | Phecode Y | Phecode Z | Phecode W
|==|Note|==|
{clinical note here}

<|eot_id|>
<|start_header_id|>assistant<|end_header_id|>
|==|Explanation|==|
{CoT}
|==|Response|==|
{label}
```

**Supplementary Figure 4**. **Structured Prompt Template for Clinical Note Summarization.** This figure presents a structured prompt template guiding an LLM in summarizing clinical notes accurately. It includes a **system header** that instructs the model to preserve the integrity of information, a **user header** detailing key categories such as demographics, medical history, symptoms, diagnostic tests, and assessments, and an **assistant header** defining the structured summary output. This approach ensures comprehensive, high-fidelity summarization for downstream genetic assessments.

```
<|begin_of_text|>
<|start_header_id|>system<|end_header_id|>
You are a genetic counselor tasked with summarizing medical notes with the highest level of detail.
Your primary objective is to capture and convey key information, ensuring the summary closely
reflects the original notes. Whenever possible, use the original wording and sentences from the
notes to preserve the accuracy and integrity of the information. Do not generate random details.
<|eot_id|>
<|start_header_id|>user<|end_header_id|>
In order to conduct a comprehensive genetic assessment, please provide as much detail as possible
about the following information:
Patient Demographics: Age, gender, ethnicity, and any relevant demographic factors.
Presenting Complaints: Describe the primary complaints or concerns, with a timeline of when
symptoms started.
Previous Assessments: Describe any past genetic evaluations or screenings, including the results
and any relevant interpretations.
Family History: Include any known instances of genetic or hereditary conditions in the family,
specifying relationships, affected members, and known genetic mutations.
Medical History: Provide a detailed personal medical history, including known medical conditions,
previous diagnoses, and treatments received.
Symptoms and Signs: List all symptoms and physical signs observed, noting their onset, duration,
and progression.
Diagnostic Tests (where available): Share results and findings from all diagnostic tests performed
descibed in the note such as: Physical examinations (if available), Blood tests (e.g., CBC,
biochemical markers), Biochemical tests (e.g., metabolic panels, enzyme activity tests), Imaging
studies (e.g., MRI, CT, ultrasound), Audiological or visual assessments, and Genetic testing
results (e.g., karyotyping, exome or genome sequencing).
Current Assessment: Provide complete ongoing evaluations or preliminary diagnoses. This is the most
important information for downstream genetic assessment.
Environmental and Lifestyle Factors: Describe relevant environmental exposures, lifestyle habits,
or occupational risks that may influence genetic predispositions.
Future Directions: Mention planned tests, referrals, or interventions that may help refine or
confirm a diagnosis.
Additional Details: Add any other context or information that may assist in making an accurate
genetic assessment.
Input:
|==|Note|==|
{clinical note here}

<|eot_id|>
<|start_header_id|>assistant<|end_header_id|>
|==|Response|==|
{Summary}
```

**Supplementary Figure 5**: This figure shows **RareDAI-Llama-3.1-8B** (ICD/CoT on summarized notes) response for the patient used for genome sequencing testing case in Figure 4. **RareDAI-Llama-3.1-8B** correctly recommended a genome sequencing.

|==|Explanation|==|
Based on the provided clinical note, I strongly recommend **genome sequencing** as the best choice for this patient. Here's a detailed, expert, and logical step-by-step reasoning explanation for why genome sequencing is strongly preferable to gene panel in this case:

1. **Congenital abnormalities or developmental disorders**: Although the patient is [PATIENT AGE] and has poor weight gain, there are no clear indications of congenital abnormalities or developmental disorders. However, the patient's symptoms and signs, such as mild respiratory distress after birth and trouble latching on to the breast, may be related to an underlying genetic condition.

2. **Multiple genes or complex phenotype**: The patient's condition involves multiple symptoms and signs, including poor weight gain, mild respiratory distress after birth, and trouble latching on to the breast. These symptoms may be related to a complex phenotype that cannot be confidently explained by a single-gene or targeted panel approach.

3. **Distinctive clinical features, biochemical findings, or imaging results**: Although the patient has some distinctive clinical features, such as thin appearance and low vital signs, the biochemical findings and imaging results are normal. This lack of clear biochemical or imaging evidence suggests that a targeted gene panel may not be the most efficient approach.

4. **Family history**: The patient's sibling has jaundice, but there is no clear indication of a specific genetic disorder based on family history. Therefore, starting with targeted testing or a gene panel may not be the most appropriate approach.

5. **Prior genetic testing or diagnostic tools**: The patient has not undergone prior genetic testing, and the results of the previous diagnostic tools, such as CBC and BMP, are not conclusive. This lack of clear diagnostic evidence suggests that genome sequencing may be a more appropriate approach.

6. **Urgent care setting**: Although the patient is not in an urgent care setting, such as the NICU or ICU, the need for rapid results for clinical management is still present. Genome sequencing can provide a quick evaluation of most protein-coding genes at once, making it a suitable choice.

7. **Cost or accessibility concerns**: Although cost or accessibility concerns may be a limitation, the patient's symptoms and signs, such as poor weight gain and mild respiratory distress after birth, justify the use of genome sequencing as a first-tier test.

In conclusion, based on the provided clinical note, **genome sequencing** is the most suitable choice for this patient. The patient's symptoms and signs, lack of clear biochemical or imaging evidence, and the need for rapid results for clinical management justify the use of genome sequencing as a first-tier test.
|==|Response|==|
**genome sequencing**

**Supplementary Figure 6**: This figure shows **ChatGPT**'s response for the patient used for genome sequencing testing case in Figure 4. **ChatGPT** incorrectly recommended a gene panel because it ignores ACMG first-tier recommendations for nonspecific, severe growth failure by favoring a narrow panel and restricts testing to selected malabsorption/metabolic genes, increasing risk of missed diagnoses across the broad differential.

Given the current presentation, I recommend starting with a **targeted gene panel** focused on failure-to-thrive/infantile feeding difficulty and malabsorption/metabolic etiologies (e.g., panels that include genes related to congenital malabsorption, pancreatic insufficiency, cystic fibrosis, inborn errors of metabolism, and endocrine/growth pathways).

Why this test:

- The patient has a specific, dominant clinical feature (poor weight gain/failure to thrive) with otherwise unremarkable exam, normal newborn screen, and limited systemic findings. This fits a scenario where a curated multi-gene panel aligned to the phenotype is efficient and cost-effective.
- Panels are faster and less expensive than exome/genome sequencing and can capture the most relevant, high-yield genes for failure to thrive and malabsorption (e.g., CFTR, SLC26A3, DGAT1, PCSK1, SLC5A1, congenital lactase deficiency genes, genes for congenital enteropathies, pancreatic insufficiency, and selected metabolic genes).
- Current clinical suspicion prioritizes nutritional intake issues with genetic/metabolic causes considered less likely; thus, a panel balances yield and turnaround while broader sequencing can be reserved if the panel is negative or if additional nonspecific/multisystem features emerge.

When to escalate to exome/genome:

- If the gene panel is non-diagnostic and the patient develops additional nonspecific or multisystem concerns, or if clinical evaluation remains inconclusive, proceed to whole exome or whole genome sequencing. This aligns with ACMG guidance to use exome/genome when panel/single-gene testing is unrevealing or the genetic cause is unclear.

**Supplementary Figure 7**. This figure shows **Llama-3.1-8B base model's** response for the patient used for genome sequencing testing case in Figure 4. **Llama** model incorrectly recommended a gene panel because it misapplies ACMG guidance by favoring a panel despite a nonspecific, severe growth failure presentation where genome/exome is recommended first-tier. It repeats unsupported assumptions (e.g., "distinctive features," family-history-driven testing, newborn screen sufficiency) and introduces contradictions.

Based on the provided input, I recommend '**gene panel**' as the genetic testing approach for the patient. Here's a detailed explanation:

**1.** The patient is presenting with congenital abnormalities or developmental disorders, specifically failure to thrive and possible malabsorption syndrome. According to ACMG guidelines, exome or genome sequencing should be considered as a first or second-tier test for these conditions to provide a comprehensive diagnostic approach. However, given the patient's age and the fact that the patient has already undergone a normal newborn screen, a targeted approach may be more efficient.

**2.** The patient's condition or suspected genetic disorder involves multiple genes or a complex phenotype that cannot be confidently explained by a single-gene or targeted panel approach. ACMG guidelines support the use of exome or genome sequencing for broader evaluation and potential reanalysis over time. However, given the patient's age and the fact that the patient has already undergone a normal newborn screen, a targeted approach may be more efficient.

**3.** The patient has distinctive clinical features, biochemical findings, or imaging results that suggest a particular genetic condition or set of conditions. The patient's symptoms, such as poor weight gain, mild respiratory distress after birth, and trouble latching on to the breast, suggest a possible metabolic or genetic etiology. A targeted gene panel may be the most efficient approach to identify causative variants.

**4.** The patient has a high likelihood of a specific genetic disorder based on family history. The patient's sibling has jaundice, which may indicate a possible genetic disorder. ACMG guidelines recommend starting with targeted testing or a gene panel to efficiently identify causative variants.

**5.** The patient has undergone prior genetic testing or any diagnostic tools, and the results were inconclusive or insufficient for diagnosis. The patient has already undergone a normal newborn screen, which did not provide a diagnosis. A targeted gene panel may be the first step to identify causative variants.

**6.** The patient is not in an urgent care setting, such as the NICU or ICU, requiring rapid results for clinical management. Therefore, rapid exome or genome sequencing is not necessary.

**7.** There are no cost or accessibility concerns that might limit the use of exome or genome sequencing as a first-tier test. However, given the patient's age and the fact that the patient has already undergone a normal newborn screen, a targeted approach may be more efficient.

Based on these factors, a **targeted gene panel** may be the most efficient approach to identify causative variants and provide a diagnosis for the patient.

**Supplementary Figure 8. Confusion matrices of RareDAI model recommendations stratified by clinician-validated versus hallucinated chain-of-thought (CoT) for 30 random testing examples.**

**A. RareDAI-Qwen3-8B**

| | Appropriate Recommendation | Inappropriate Recommendation |
|---|---|---|
| Validated CoT | 23 | 2 |
| Hallucinated CoT | 2 | 3 |

**B. RareDAI-Llama3.1-8B**

| | Appropriate Recommendation | Inappropriate Recommendation |
|---|---|---|
| Validated CoT | 23 | 2 |
| Hallucinated CoT | 1 | 4 |

**Supplementary Figure 9**: **Pairwise McNemar test results across models.** Heatmap of McNemar p-values comparing model prediction differences, showing no significant differences among models trained on the same note type (full vs. full, summarized vs. summarized) and significant differences primarily between full-note and summarized-note models.

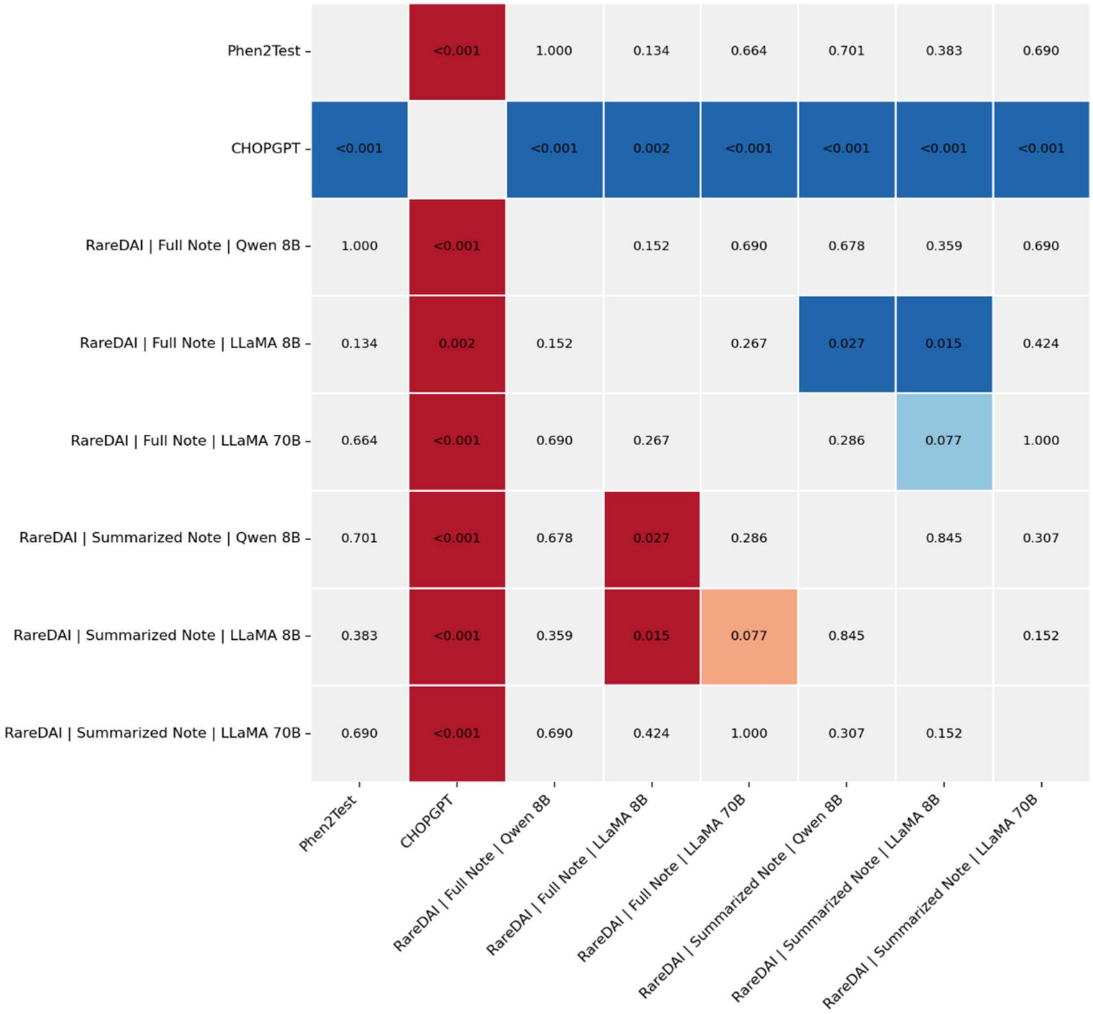

**Supplementary Figure 10**: **Feature importance in Phen2Test Random Forest.** Clinical note text, sensorineural hearing loss, autism, developmental delay, and age are the top contributors to genetic test recommendation in the CHOP cohort.

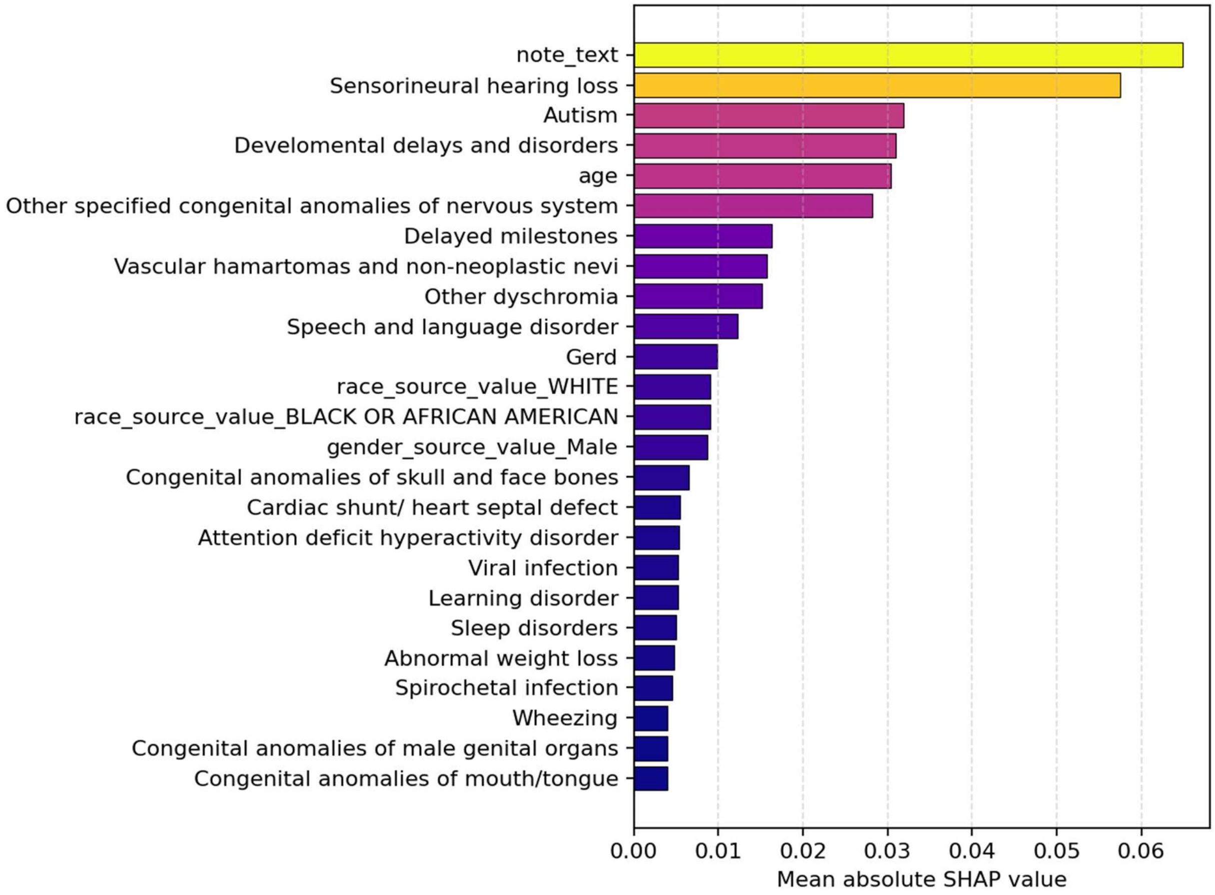

**Supplementary Figure 11**. Performance comparison between zero-shot and few-shot CHOPGPT under different evaluation criteria on 30 manual testing examples. This figure reports the GT-Accuracy (Ground Truth) and Adjusted-Accuracy of CHOPGPT (after clinical expert reviews) under zero-shot and few-shot prompting settings. Few-shot prompting substantially improves performance compared to zero-shot, particularly under the adjusted evaluation, indicating that in-context examples enhance the model's robustness and alignment with clinically adjusted ground truth.

| | GT-Accuracy | Adjusted-Accuracy |
|---|---|---|
| **Zero-shot CHOGPT** | 0.53 | 0.45 |
| **Few-shot CHOPGPT** | 0.63 | 0.83 |

**Supplementary Figure 12**: The figure below shows that the reduction in precision (or quantization) causes information loss, leading to lower scores in all metrics, regardless of fine-tuning or base models. These models trained on long clinical notes along with HPO/ICD/CoT.

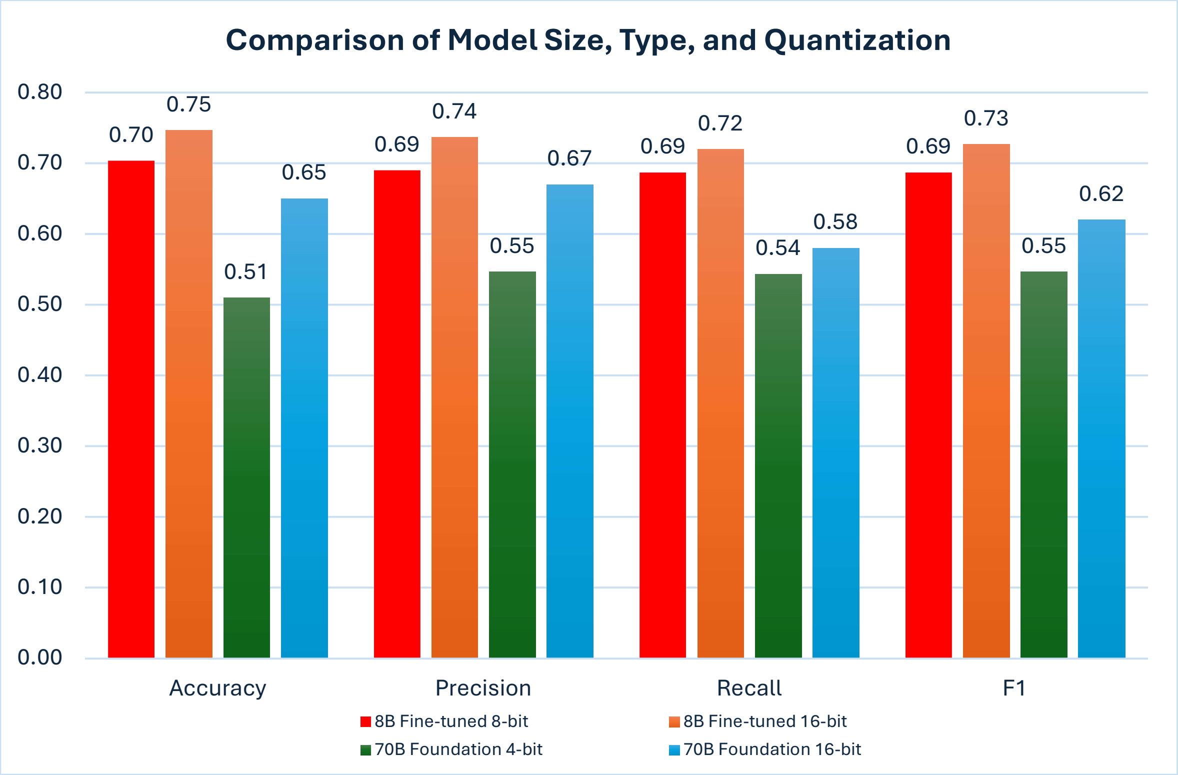

**Supplementary Table 1**: The table below shows the performances of base Llama-3.1-8B (half precision) and 70B (4bit and 16bit) models on the CHOP test dataset. Even without fine-tuning, the 70B model with CoT questions outperforms the others, demonstrating the effectiveness of CoT questions.

| Model | Base (8B) | | Base (70B) | | Base (70B) | |
|---|---|---|---|---|---|---|
| Precision | 16bit | | 4bit | | 16bit | |
| Prompt | Without Questions | With Questions | Without Questions | **With Questions** | Without Questions | **With Questions** |
| Accuracy | 0.64 | 0.63 | 0.51 | **0.52** | 0.65 | **0.65** |
| Precision | 0.63 | 0.61 | 0.55 | **0.58** | 0.67 | **0.76** |
| Recall | 0.60 | 0.56 | 0.54 | **0.57** | 0.58 | **0.56** |
| F1 | 0.61 | 0.58 | 0.55 | **0.58** | 0.62 | **0.65** |
